# B and T cell responses to pre-erythrocytic R21/Matrix-M and blood-stage RH5.1/Matrix-M malaria vaccines in endemic settings

**DOI:** 10.1101/2025.11.11.25339978

**Authors:** Caroline K Bundi, Olivia Muñoz, Elizabeth Kibwana, Duncan Bellamy, Lisa Stockdale, Jordan R Barrett, Kirsty McHugh, Ivanny Mtaka, Wilmina Kalinga, Domtila Kimani, Lydia Nyamako, Kelvias Keter, Rodney Ogwang, Adrian VS Hill, Philip Bejon, Mainga Hamaluba, Sarah E Silk, Angela M Minassian, Simon J Draper, Ally I Olotu, Melissa Kapulu, Katie J Ewer, Carolyn M Nielsen

## Abstract

There is great interest in combining the licensed pre-erythrocytic malaria vaccine R21/Matrix-M^®^ and the blood-stage candidate vaccine RH5.1/Matrix-M^®^ to maximise efficacy against *Plasmodium falciparum* malaria. As protection is understood to be antibody-mediated for both vaccines, the success of a multi-stage vaccination strategy will be aided by understanding factors that impact the cellular drivers of humoral immunity, e.g. dosing regimen, exposure to malaria, and age. Prior analyses of adult vaccinees showed higher RH5-specific memory B cell responses with delayed fractional versus monthly booster dosing (50-50-10µg at 0-1-6months and 10-10-10µg at 0-1-2months, respectively), but the B cell impact of delayed booster regimens has not yet been evaluated in the target paediatric population. There have also not yet been NANP-specific B cell immunogenicity analyses with R21/Matrix-M. Here, pre- and post-vaccination PBMC from two Phase 1b clinical trials with R21/Matrix-M^®^ (NCT03580824; Kenya) and RH5.1/Matrix-M^®^ (NCT04318002; Tanzania) were analysed by flow cytometry for antigen-specific memory B cell and total (R21) or antigen-specific (RH5) circulating Tfh (cTfh) cell responses. For R21, higher frequencies of NANP-specific activated memory IgG⁺ B cells were detected in infants versus adults, and also with a higher Matrix-M^®^ dose (50µg versus 25µg). Both NANP-specific IgG⁺ memory B cells and total cTfh cells correlated with anti-NANP serum IgG. For RH5.1, higher previous malaria exposure was associated with increased RH5-specific cTfh and cTfh2 cell frequencies. RH5-specific IgG⁺ memory B cell responses were greatest with delayed booster dosing (10-10-10µg at 0-1-6months) compared to monthly or delayed fractional regimens, and correlated with both peak and late time point serum antibody (2-years after final vaccination). Interestingly, while late time point anti-RH5.1 serum IgG concentrations in delayed booster children vaccinees matched previous reports with RH5.1/AS01_B_ delayed fractional dosing in adults (NCT02927145; UK), IgG durability and relationship to RH5-specific B cells differed between these two cohorts. While our conclusions would be strengthened by further analyses in larger clinical trials, our data indicate that age, adjuvant dose, vaccine dose, and timing of final booster vaccination impact antigen-specific cellular responses to malaria vaccines and could thus inform the development of next-generation multi-stage malaria vaccination strategies.

## Introduction

Immunisation is a key pillar in global health, saving millions of lives each year (WHO, 2023b). Vaccines play a vital role in controlling, eliminating, and potentially eradicating diseases, with smallpox serving as a powerful example (Shchelkunova and Shchelkunov, 2017). Vaccines work by generating immune memory, enabling a rapid response to future pathogen exposure through memory B cells and T cells (Palm and Henry, 2019). For malaria, the rapid decline of vaccine-induced immunity remains a major challenge (Minassian *et al*., 2021; Datoo *et al*., 2022; Nielsen *et al*., 2023; Dicko *et al*., 2024), making it crucial to understand how existing malaria vaccines induce and sustain a strong immune response.

Recent advances in malaria vaccine development have been significant, particularly with the licensing and WHO recommendation of RTS,S/AS01 and R21/Matrix-M (WHO, 2021, 2023a). These vaccines target the pre-erythrocytic stage of *Plasmodium falciparum*, aiming to prevent infection by inhibiting the parasite’s development immediately after mosquito inoculation. Both are based on the circumsporozoite protein (CSP) fused to the hepatitis B surface antigen (Collins *et al*., 2017; Laurens, 2020), and both show correlation between IgG antibody levels to the four amino acid repeat region (NANP) and clinical efficacy (White *et al*., 2014; Datoo *et al*., 2024). In contrast, blood-stage vaccines target the asexual parasite forms responsible for clinical disease. Among the candidates in development, *P. falciparum* reticulocyte-binding protein homolog 5 (RH5) is the leading blood-stage antigen. The RH5.1/Matrix-M^®^ formulation has shown encouraging results in a Phase 1b trial in Tanzania and is currently under evaluation in a Phase 2b trial in Burkina Faso (Silk *et al*., 2024; Natama *et al*., 2025). Notably, protection from both R21 and RH5-based vaccines is primarily antibody-mediated, highlighting the importance of understanding B cell and T follicular helper (Tfh) cell responses in driving robust and durable humoral immunity.

Data from the RH5.1 programme indicate stronger humoral responses in children compared to adults, particularly with a delayed booster schedule (0-1-6 months, 10-10-10µg antigen), which outperformed both monthly (0-1-2 months, 10-10-10µg antigen) and delayed fractional (antigen) dosing regimens (0-1-6 months, 50-50-10µg) in eliciting antibody responses (Silk *et al*., 2024). Interim efficacy results from the ongoing Phase 2b trial show 55% efficacy at 6 months with the delayed regimen, compared to 40% with the monthly schedule (Natama *et al*., 2025), with 12-month efficacy data forthcoming. The trial is also testing the combination of R21 and RH5.1 vaccines in children; these first proof-of-concept multi-stage vaccine efficacy results are eagerly awaited. Additional trials are also underway, including one combining RH5.1 and R78C (a fusion protein of the C-terminal EGF-like domains of RIPR with CyRPA, both components of the invasion complex used in RH5-mediated invasion of red blood cells (Williams *et al*., 2024) in adults and children in the UK and Tanzania (NCT05385471; PACTR202407803870883), and another evaluating delayed versus delayed fractional dosing regimens of RH5.1/Matrix-M^®^ in UK adults (NCT06141057). These multi-stage / multi-antigen vaccine studies mark a promising step toward broader and more effective malaria control.

Varying vaccination booster schedules and antigen doses have also been shown to influence the frequencies of the vaccine-induced memory B cells in other programmes (Pallikkuth *et al*., 2020; Nicolas *et al*., 2023; Nielsen *et al*., 2023; Barrett *et al*., 2024; Hodgson *et al*., 2025). Specifically, a delayed fractional vaccine antigen dose has been associated with higher frequencies of memory B cells compared to monthly dosing (Pallikkuth *et al*., 2020; Nielsen *et al*., 2023; Barrett *et al*., 2024). The vaccination platform has also been shown to influence the RH5-specific circulating T follicular helper (cTfh) cell response in RH5 vaccinees, with the RH5.1/AS01_B_ protein/adjuvant platform inducing higher responses compared to RH5 delivered by a heterologous viral vector platform (Nielsen *et al*., 2021).

Various other factors influencing malaria vaccine-induced humoral responses have also been proposed (Zimmermann and Curtis, 2019; van Dorst *et al*., 2024). These include: variation of the vaccination regimens (antigen and adjuvant dose), age of the vaccinee, and impact of previous exposure to malaria. Published data across R21 and RH5.1 clinical trials suggest all of these factors may be important (Datoo *et al*., 2021; Minassian *et al*., 2021; Nielsen *et al*., 2021, 2023; Sang *et al*., 2023; Silk *et al*., 2023, 2024; Barrett *et al*., 2024; Bundi *et al*., 2025; Venkatraman *et al*., 2025). For example, in a Phase 1 trial, R21 showed higher anti-NANP and anti-C-terminus IgG antibodies in infants compared to both older children (18-36mo) and adults (Sang *et al*., 2023; Bundi *et al*., 2025), and in a Phase 3 trial, younger children aged 5-17 months had higher titres compared with children aged 18-36 months (Datoo *et al*., 2024). A Phase 2 trial with n=150 per group showed increased efficacy using a higher Matrix-M^®^ adjuvant dose (50µg) compared with a half-dose (25µg) when the R21 dose was held the same (5µg), although the mechanism behind this finding remains unclear (Datoo *et al*., 2021, 2022). In a Phase 3 trial, R21/Matrix-M^®^ showed efficacy in areas with seasonal (75%) and perennial (68%) malaria transmission with a dose of 5μg R21/50μg Matrix-M^®^ (Datoo *et al*., 2024). NANP-specific B cell responses have not previously been analysed in these contexts.

While B cells are the cellular source of antibodies, Tfh cells are essential for B cell support in germinal centres (GCs), aiding the production of memory B cells and plasma cells (De Silva and Klein, 2015). Analogous to total CD4+ T cells, Tfh can be classified into Tfh1 (CXCR3+ CCR6), Tfh2 (CXCR3-CCR6-), and Tfh17 (CXCR3-CCR6+) (Crotty, 2014; Qi, 2016). Secondary lymphoid tissues are ideal for studying Tfh cells, but ethical and logistical limitations often hinder sampling for research. However, cTfh cells (expressing CXCR5 -/+ programmed cell death protein 1 (PD1)) in peripheral blood share similar phenotypic and functional profiles with GC Tfh cells and are thus useful proxies for studying GC responses (Locci *et al*., 2013; Brenna *et al*., 2020). The important role played by Tfh in vaccination has been demonstrated in influenza, (Koutsakos *et al*., 2018), HIV (Locci *et al*., 2013), SARS-CoV-2 (Mudd *et al*., 2022), and yellow fever (Huber *et al*., 2020) vaccines, among others. Frequencies of vaccine-specific cTfh cells have also been shown to correlate with serum antibodies in RTS,S/AS01, and earlier RH5.1/AS01_B_ clinical trials (Bowyer *et al*., 2018; Hill *et al*., 2019; Pallikkuth *et al*., 2020; Nielsen *et al*., 2021).

Despite the above evidence, R21- and RH5.1-specific B and cTfh cell responses in the target populations in malaria-endemic regions remain unexplored. In this study, we sought to address these data gaps and evaluated the effect of vaccination regimens (vaccination dose, adjuvant dose, vaccination schedule), age, and malaria exposure on RH5.1/Matrix-M^®^ and, separately, R21/Matrix-M^®^.

## Methods

### Clinical trials

This study was nested within two independent clinical trials: R21/Matrix-M^®^ (NCT03580824; 2019-2022) and RH5.1/Matrix-M^®^ (NCT04318002; 2021-2023). We additionally compared a subset of the RH5.1/Matrix-M^®^ (NCT04318002) read-outs to previously published data with an RH5.1/AS01_B_ clinical trial in a malaria-naïve UK adult population (NCT02927145; 2016-2019). Sample sizes for the analyses were determined by sample availability and are summarised in the Supplementary Table 1. The group sizes shown in Tables 1-3 represent the maximum sample size as defined by trial enrolment.

**Table 1:** R21/Matrix-M^®^ - Kenya (NCT03580824) groups included in this study.

| Group (Age) | Number of participants | Month 0 (V1) | Month 1 (V2) | Month 2 (V3) |
| --- | --- | --- | --- | --- |
| Group 1A/B<br>Adults (18-45 years) | 18 | 10µg R21<br>/50µg Matrix-M<sup>®</sup> | 10µg R21<br>/50µg Matrix-M<sup>®</sup> | 10µg R21<br>/50µg Matrix-M<sup>®</sup> |
| Group 3A/C<br>Infants (5-11 months) | 18 | 5µg R21<br>/25µg Matrix-M<sup>®</sup> | 5µg R21<br>/25µg Matrix-M<sup>®</sup> | 5µg R21<br>/25µg Matrix-M<sup>®</sup> |
| Group 3B/D<br>Infants (5-11 months) | 18 | 10µg R21<br>/50µg Matrix-M<sup>®</sup> | 10µg R21<br>/50µg Matrix-M<sup>®</sup> | 10µg R21<br>/50µg Matrix-M<sup>®</sup> |
| Group 3E<br>Infants (5-11 months) | 15 | 5µg R21<br>/50µg Matrix-M<sup>®</sup> | 5µg R21<br>/50µg Matrix-M<sup>®</sup> | 5µg R21<br>/50µg Matrix-M<sup>®</sup> |

V1: Vaccination 1, V2: Vaccination 2, V3: Vaccination 3.

**Table 2:** RH5.1/Matrix-M^®^ - Tanzania (NCT04318002) groups included in this study.

| Group (Age) | Number of participants | Malaria Pre-exposure status* | Month 0 (V1) | Month 1 (V2) | Month 2 (V3) (monthly regimen) | Month 6 (V3) (delayed regimen) |
| --- | --- | --- | --- | --- | --- | --- |
| Group 1A adults (18-45 years) | 6 | Low | 10µg RH5.1 /50µg Matrix-M<sup>®</sup> | 10µg RH5.1 /50µg Matrix-M<sup>®</sup> | 10µg RH5.1 /50µg Matrix-M<sup>®</sup> |  |
| Group 1B adults (18-45 years) | 6 | Low | 50µg RH5.1 /50µg Matrix-M<sup>®</sup> | 50µg RH5.1 /50µg Matrix-M<sup>®</sup> |  | 10µg RH5.1 /50µg Matrix-M<sup>®</sup> |
| Group 2A children (5-17 months) | 12 | Low | 10µg RH5.1 /50µg Matrix-M<sup>®</sup> | 10µg RH5.1 /50µg Matrix-M<sup>®</sup> | 10µg RH5.1 /50µg Matrix-M<sup>®</sup> |  |
| Group 2B children (5-17 months) | 12 | Low | 10µg RH5.1 /50µg Matrix-M<sup>®</sup> | 10µg RH5.1 /50µg Matrix-M<sup>®</sup> |  | 10µg RH5.1 /50µg Matrix-M<sup>®</sup> |
| Group 2C children (5-17 months) | 12 | Low | 50µg RH5.1 /50µg Matrix-M<sup>®</sup> | 50µg RH5.1 /50µg Matrix-M<sup>®</sup> |  | 10µg RH5.1 /50µg Matrix-M<sup>®</sup> |
| Group 2D children (5-17 months) | 12 | High | 50µg RH5.1 /50µg Matrix-M<sup>®</sup> | 50µg RH5.1 /50µg Matrix-M<sup>®</sup> |  | 10µg RH5.1 /50µg Matrix-M<sup>®</sup> |

V1: Vaccination 1, V2: Vaccination 2, V3: Vaccination 3. Note that cellular analyses of the adult groups have been published elsewhere (Barrett *et al.*, 2024).
\* Before the start of the trial, a serological assay testing IgG binding to *P. falciparum* schizont lysate was established using sera from children living in urban and rural wards within Bagamoyo District to define relative “low” versus “high” thresholds of malaria pre-exposure relevant to this study population. These thresholds were used to assign children to groups on the basis of malaria pre-exposure in this study (method described further in (Silk *et al.*, 2024). We note that the “low” versus “high” exposure statuses used in this trial are contextual and relative to the specific study population. All vaccinees are likely lower exposure than if the trial had been conducted in a higher endemicity setting such as e.g. Burkina Faso.

**Table 3:** RH5.1/AS01_B_ – UK (NCT02927145) groups included in this study.

| Group (Age) | Number of participants | Month 0 (V1) | Month 1 (V2) | Month 2 (V3)<br>(monthly regimen) | Month 6 (V3)<br>(delayed regimen) |
| --- | --- | --- | --- | --- | --- |
| Group 2<br>Adults (18-45 years) | 12 | 10µg RH5.1<br>/0.5mL AS01<sub>B</sub> | 10µg RH5.1<br>/0.5mL AS01<sub>B</sub> | 10µg RH5.1<br>/0.5mL AS01<sub>B</sub> |  |
| Group 3<br>Adults (18-45 years) | 12 | 50µg RH5.1<br>/0.5mL AS01<sub>B</sub> | 50µg RH5.1<br>/0.5mL AS01<sub>B</sub> |  | 10µg RH5.1<br>/0.5mL AS01<sub>B</sub> |

V1: Vaccination 1, V2: Vaccination 2, V3: Vaccination 3.

The R21/Matrix-M^®^ “VAC073” trial (NCT03580824) was a Phase 1b, open-label, age de-escalation, dose-escalation trial conducted at KEMRI, Kilifi, Kenya, involving adults (18–45 years), children (1–5 years; not included in the analyses presented here), and infants (5-11 months). Vaccines were administered on a 0-1-2 month regimen, with groups varying in the dose of R21 antigen and/or Matrix-M^®^ adjuvant (Table 1). Full details of the clinical trial can be found in the primary trial publication, including analyses of 1-5-year olds and antibody responses following further booster doses which were outside the scope of this study (Sang *et al*., 2023; Bundi *et al*., 2025).

The RH5.1/Matrix-M^®^ “VAC080” trial (NCT04318002) was also a Phase 1b, open-label, age de-escalation dose-escalation trial that was conducted at Ifakara Health Institute, Bagamoyo, Tanzania. The trial recruited healthy adults (18-45 years); not included in the analyses presented here (Barrett *et al*., 2024), and children (5-17 months) (Silk *et al*., 2024). Vaccines were administered on a 0-1-2 month “monthly” or 0-1-6 month “delayed” regimen, with delayed regimen groups varying in terms of the vaccine antigen dose of the final booster (“delayed” versus “delayed fractional”) and pre-exposure to malaria infection (Table 2). Full details of the clinical trial, including analyses of adult participants, can be found in the primary trial publication (Silk *et al*., 2024).

To note, in the interest of consistency we use the age group nomenclature as per primary trial publications, i.e. 5-11mo “infants” in the R21/Matrix-M^®^ trial (NCT03580824) (Sang *et al*., 2023) and 5-17mo “children” in the RH5.1/Matrix-M^®^ trial (NCT04318002) (Silk *et al*., 2024). We do not seek to draw any comparisons between these two overlapping age groups in this study.

The RH5.1/AS01_B_ “VAC063” trial (NCT02927145) was a Phase 1/2a trial conducted in malaria-naïve adults in the UK to determine the safety, immunogenicity, and efficacy of RH5.1/AS01_B_ against blood-stage controlled human malaria infection (CHMI) model (Minassian *et al*., 2021). Vaccines were administered on a 0-1-2 “monthly” or 0-1-6 month “delayed fractional regimen” (Table 3). Full details of the clinical trial, including groups not included in this study, can be found in the primary trial publication (Minassian *et al*., 2021).

All three trials were conducted according to the principles of the Declaration of Helsinki 2013 and Good Clinical Practice. For the RH5.1/Matrix-M^®^ trial (NCT04318002), guardians of the child participants signed or thumb-printed an informed consent form at the in-person screening visit, and consent was verified verbally before each vaccination. For the RH5.1/AS01_B_ trial (NCT02927145), all participants signed written consent forms, and consent was checked before each vaccination and prior to CHMI. For the R21/Matrix-M^®^ trial (NCT03580824) written informed consent for participation in this trial was provided by adult volunteers, whilst the children and infant participants’ parents or legal guardians provided consent for participation in this trial, which allowed the use of the sample and data for the primary trial objective and future exploratory analysis.

### RH5-specific and NANP-specific B cell flow cytometry

Cryopreserved PBMCs from the RH5.1/Matrix-M^®^ and R21/Matrix-M^®^ vaccine trials were thawed using pre-warmed R10 media (RPMI [R0883, Sigma] supplemented with 10% heat-inactivated FCS [60923, Biosera], 100U/ml penicillin/0.1mg/mL streptomycin [P0781, Sigma], and 2mM L-glutamine [G7513, Sigma]). Thawed PBMC were washed twice and rested in R10 for 1h at 37°C with benzonase (70664-10KUN, EMD Millipore). The PBMCs were enriched for B cells using the Human Pan-B cell Enrichment Kit [19554, StemCell]. Enriched B cells were then stained with viability dye FVS780 (565388, BD Biosciences), followed by incubation with Trustain (422302, Biolegend) to block non-specific Fc receptor staining. Subsequent surface staining of B cells was performed with anti-human CD19-BV786 (563325, BD Biosciences), IgG-BB515 (564581, BD Biosciences), IgM-BV605 (562977, BD Biosciences), CD27-PE-Cy7 (560609, BD Biosciences), CD21-BV711 (563163, BD Biosciences) IgA-PerCP-Vio700 (130-113-478, Miltenyi), CD38-BV480 (566137, BD Biosciences), and CD138-APC-R700 (566050, BD Biosciences). Two fluorophore-conjugated NANP or RH5 probes, for R21/Matrix-M^®^ and RH5.1/Matrix-M^®^ trial samples respectively, were also included. The preparation of the RH5 probes has been published previously (Nielsen *et al*., 2023; Barrett *et al*., 2024). In brief, monobiotinylated RH5 was produced by transient co-transfection of HEK293F cells with a plasmid encoding BirA biotin ligase and a plasmid encoding a full-length RH5. The RH5 plasmid was based on ‘RH5-bio’ (a gift from Gavin Wright; University of York, York, United Kingdom; Addgene plasmid 47780; http://n2t.net/addgene:47780;RRID: Addgene_47780; (Crosnier et al., 2013). RH5-bio was modified prior to transfection to incorporate a C-tag for subsequent protein purification. The monobiotinylated NANP peptide used for probe generation was commercially obtained (CPD70698, Thinkpeptides, ProImmune). Two probes per antigen were freshly prepared for each experiment, by incubation of monobiotinylated RH5 or NANP with streptavidin-PE (S866, Invitrogen) or streptavidin-APC (17-4317-82, Biolegend) at an approximately 4:1 molar ratio to facilitate tetramer generation. Probes were then centrifuged at 13000rpm (max microfuge speed) for 10min at room temperature to remove aggregates. After surface staining, enriched B cells were permeabilised and fixed with Transcription Factor Buffer Set (562574, BD Biosciences) to enable intracellular staining. The fixed cells were stained with anti-human Ki67-BV650 (563757, BD Biosciences; data not shown) and probes at 1/10 dilution as compared to concentrations used for surface staining. The cells were washed and stored at 4°C until acquisition.

B cells were acquired on a Fortessa X20 flow cytometer with FACSDiva8.0 (BD Biosciences). Data were analysed using FlowJo (v10.8; Tree Star Inc.). Samples were excluded from analysis if there were < 50 cells in the parent population. NANP-or RH5-specific IgG+ B cells were identified as RH5/APC+RH5/PE+ or NANP/APC+NANP/PE+ cells within various live memory B cell populations as shown in Supplementary Figure 1.

### *Ex vivo* cTfh cell assay

Technical issues with our CSP stimulation meant we were unable to investigate frequencies of CSP-specific cTfh cells within total memory cTfh cells from available PBMC in the R21/Matrix-M^®^ trial (NCT03580824). Instead, we used a different read-out as a proxy for vaccine-induced cTfh cell responses 7 days after the final vaccination in the primary three-dose series (V3+7), total CXCR5+PD1+ cTfh cells.

The *ex vivo* total cTfh assay was adapted from previously published methodology (Bowyer *et al*., 2018). In brief, cryopreserved PBMC from the R21/Matrix-M^®^ clinical trial (NCT03580824) were thawed, washed twice in R10 as above, and then rested for a further 3h at 37°C. PBMC were then stained with viability dye Live/Dead Aqua (L34957, ThermoFisher) and washed before surface staining with anti-human PD1-BV650 (329950, Biolegend), CD45RA-eF450 (304123, Biolegend), CXCR5-FITC (356913, Biolegend), CCR6-PE (G034E3, G034E3), CD4-APC-eF780 (47-0049-42, eBioscience), CD3-AF700 (56-0038-82, eBioscience) and CXCR3-APC (353708, Biolegend) at 37°C. Cells were fixed with 1% paraformaldehyde (43368, Alfa Aesar) and stored at 4°C until acquisition on the same day.

The cells were acquired on a Fortessa X20 flow cytometer with FACSDiva8.0 (BD Biosciences). Data were analysed using FlowJo (v10.8; Tree Star Inc.). Samples were excluded from analysis if there were < 50 cells in the parent population. Memory CD4+ T cells were identified as live CD4+ CD45RA- cells, and cTfh were defined as CXCR5+PD1+ within the memory CD4+ T cells as shown in Supplementary Figure 2.

### Activation Induced Marker (AIM) Assay

The AIM assay was adapted from previously published methodology(Nielsen *et al*., 2021). In brief, cryopreserved PBMC from the RH5.1/Matrix-M^®^ clinical trial (NCT04318002) were thawed as above and washed twice in R10 before resting for 3h at 37°C with benzonase (70664-10KUN, EMD Millipore). This was followed by 22h stimulation with either R10 medium alone as a negative control, 2.5μg/peptide/mL of a RH5 peptide pool (20-mer peptides spanning full-length RH5 protein, overlapping by 10 amino acids, total of 50 peptides [NeoScientific] (Payne *et al*., 2017; Nielsen *et al*., 2021) or 1μg/mL Staphylococcal enterotoxin B (SEB; S-4881, Sigma; positive control). Following stimulation, PBMC were surface stained as per the *ex vivo* cTfh cell panel above as well as anti-human CD134 (OX40)-BUV395 (743286, BD Biosciences), CD25-PE-Cy7 (335824, BD Biosciences), and CD69-PE-Cy5 (555532, BD Biosciences). Finally, the cells were washed and then fixed with 1% paraformaldehyde (43368, Alfa Aesar) and stored at 4°C until acquisition on the same day.

Samples were acquired on a Fortessa X20 flow cytometer with FACSDiva8.0 (BD Biosciences). Data were analysed using FlowJo (v10.8; Treestar). Samples were excluded from analysis if there were <50 cells in the parent population. RH5-specific cells within memory CD4+ or cTfh cells were defined as CD25+OX40+ or CD25+CD69+ as shown in Supplementary Figures 2-4. Background responses to medium alone were subtracted from RH5-specific responses in matched samples.

### IgG standardised ELISA

Standardised ELISAs were used to quantify the magnitude of the anti-NANP and anti-RH5 serum IgG pre- and post-vaccination with R21/Matrix-M^®^ and RH5.1/Matrix-M^®^ vaccines, respectively. Anti-RH5.1 total IgG ELISAs were performed against full-length RH5 protein (RH5.1) using standardised methodology as previously described (Silk *et al*., 2023, 2024). Anti-NANP total IgG antibodies were measured against a synthetic peptide comprising 6 repeats of the amino acid sequence NANP (NANP6), as previously described (Sang *et al*., 2023). In both trials, the reciprocal of the test sample dilution giving an OD_405_ of 1.0 in the standardised assay was used to assign an ELISA unit value of the standard. A standard curve and Gen5 ELISA software v3.04 (BioTek, UK) were used to convert the OD_405_ of individual test samples into AU. In RH5 vaccinees, the AU were converted into μg/mL following generation of a conversion factor by calibration-free concentration analysis as reported previously (Payne *et al*., 2017).

### Statistical Analyses

Data was analysed using GraphPad Prism version 10.1 for Windows (GraphPad Software Inc., California, USA). Comparisons between different time points/groups were done with Mann-Whitney U tests and/or a Kruskal-Wallis test with Dunn’s correction for multiple comparisons. Spearman’s rank correlation was used to test associations. All p values (or adjusted p values for tests with Dunn’s correction) are two-tailed and were considered significant at the value of < 0.05. Only significant results are annotated on figures but all statistical tests performed (including those with non-significant results) are detailed in figure legends.

## Results

### NANP-specific B cell responses to R21/Matrix-M^®^ are influenced by age and adjuvant dose

Within the R21/Matrix-M^®^ clinical trial (NCT03580824), NANP probes were used to identify circulating NANP-specific IgG+, IgM+, and IgA+ memory B cells (mBC; defined as shown in Figure 1A and Supplementary Figure 1) in PBMC samples at baseline (Day 0 (D0)) and 28 days after the final vaccination in the primary three-dose series (V3+28). The groups from NCT03580824 included in this analysis are summarised in Table 1. In brief, this comprises Group 1A/B (adults; 10µg R21/ 50µg Matrix-M^®^ at 0-1-2 months), Group 3A/C (infants; 5µg R21/ 25µg Matrix-M^®^ at 0-1-2 months), Group 3B/D (infants; 10µg R21/ 50µg Matrix-M^®^ at 0-1-2 months), and Group 3E (infants; 5µg R21/ 50µg Matrix-M^®^ at 0-1-2 months.) Significant NANP-specific responses were observed within total, resting, or activated memory IgG+, IgA+, and IgM+ B cell populations between D0 and V3+28 within all adult and infant groups, with the only exception of resting IgA+ memory B cells in Groups 3B/D and 3E, activated and resting IgM+ in Group 3B/D and 3A/C, respectively (Supplementary Figure 5). Of the different mBC subsets, the highest median responses were observed within the activated IgG+ memory subset (Figure 1B; Supplementary Figure 5).

**Figure 1.**
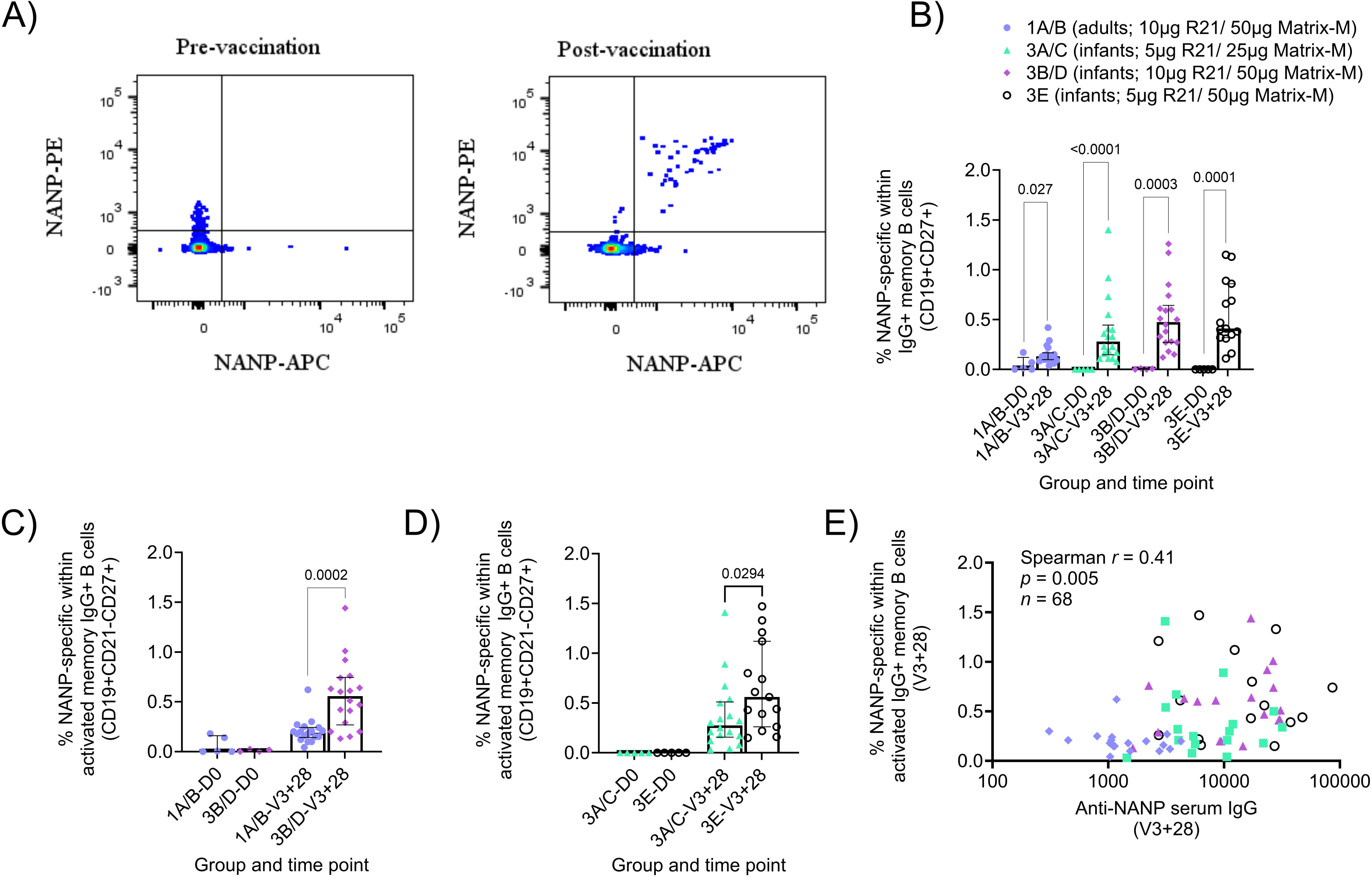
NANP-specific memory B cell responses to R21/Matrix-M^®^. Cryopreserved PBMCs from Day 0 ((D0); baseline) and V3+28 (28 days post-third vaccination (V3)) were stained *ex vivo* and analysed by flow cytometry. NANP-specific cells within total/ activated memory B cell populations were defined as shown in Figure (**A**) and Supplementary Figure 1. Frequencies were compared between (**B**) NANP-specific total IgG+ memory B cells at D0 and V3+28 within groups, (**C**) NANP-specific activated IgG+ memory B cells in adults [Group 1A/B] and infants [Group 3B/D] at D0 and V3+28, and (**D**) NANP-specific activated IgG+ memory B cells in low [Group 3A/C] or high [Group 3E] Matrix-M^®^ infants at D0 and V3+28. A Spearman correlation analysis was performed between NANP-specific activated IgG+ memory B cells and anti-NANP serum IgG at V3+28 in all groups (**E**). Post-vaccination responses were compared within groups by the Mann-Whitney test (**B**) or between groups (**C-D**). Sample sizes for all assays were based on sample availability; each point represents a single sample. Bars and error bars represent medians and interquartile ranges, respectively. Significant p values are annotated on graphs; p < 0.05 was considered significant.

In terms of differences between groups, the comparisons we were most interested in within the R21/Matrix-M^®^ trial were: Group 1A/B versus Group 3B/D (adults versus infants; same doses of R21 and Matrix-M^®^), Group 3A/C versus Group 3E (10µg versus 50µg Matrix-M^®^ in infants; same dose R21), and Group 3B/D versus Group 3E (5µg versus 10µg R21 in infants; same dose of Matrix-M^®^). Post-vaccination NANP-specific B cell frequencies were significantly higher in infants than adults (receiving the same 10μg R21/50μg Matrix-M^®^ regimen; Groups 1A/B and 3B/D) within total memory IgG+ and IgA+ B cells, as well as within activated and resting memory IgG+ subpopulations (activated memory IgG+ Figure 1C; V3+28 medians: 0.19% Group 1A/B adults, 0.56% Group 3B/D infants p = 0.0002; Supplementary Figure 6).

There were no significant differences by age within any of the NANP-specific memory IgM+ B cell populations (Supplementary Figure 6). Within the infant groups, an increase in Matrix-M^®^ dose from 25µg (Group 3A/C) to 50µg (Group 3E) with the same R21 dose was weakly associated with increased activated memory IgG+ B cells (Figure 1D; V3+28 medians: 0.28% Group 3A/C 25µg Matrix-M^®^, 0.56% Group 3E 50µg Matrix-M^®^, p = 0.0294). Increasing the R21 antigen dose showed no significant difference in the NANP-specific mBC populations (Group 3B/D versus Group 3E; data not shown). When pooling all groups analysed, frequencies of NANP-specific cells within memory IgG+ B cells correlated with matched time point V3+28 serum anti-NANP IgG (activated memory IgG+ Figure 1E; Supplementary Figure 7), but not late time point serum anti-NANP IgG titres (V3+1yr infants, V3+2yr adults; data not shown). To note, these differences were no longer significant when groups were analysed separately.

### Trend towards higher frequencies of cTfh1 and lower frequencies of cTfh17 within CXRC5+PD1+ cTfh cells in infants as compared to adults R21/Matrix-M^®^ vaccinees

While there were no significant post-vaccination differences in memory CD4+ T cells within total T cells in all groups (Figure 2A), there were weak trends to increased frequencies of CXCR5+PD1+ cTfh cells within the memory CD4+ T cell population and this was significant in adults (Figure 2B; Group 1A/B adults D0 median: 2.47% vs V3+7 median: 3.31% p = 0.0376). Few baseline samples were assayed in the infant groups; thus we were not able to meaningfully compare pre- and post-vaccination responses (Figure 2A-B).

**Figure 2.**
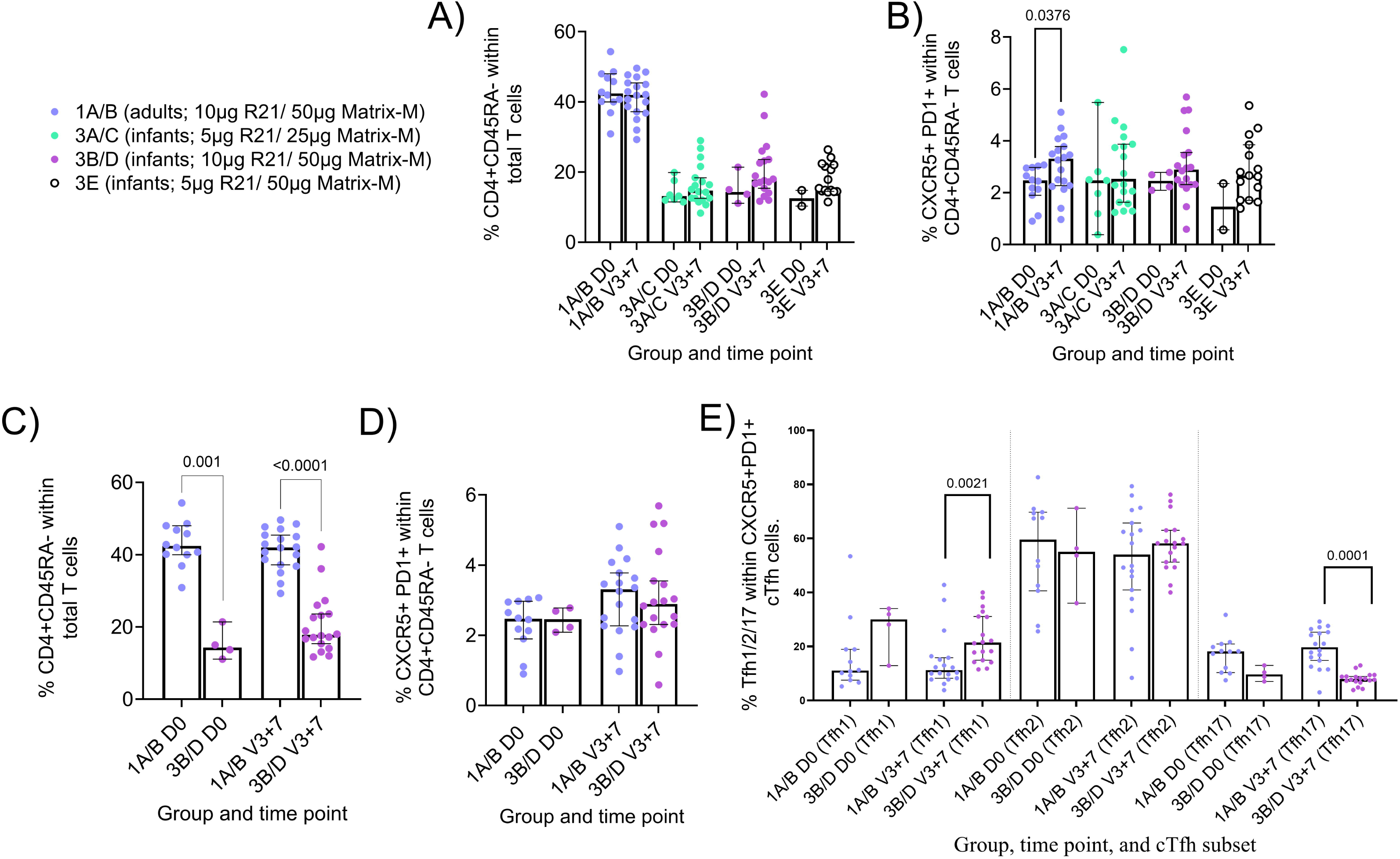
Memory CD4+ and circulating Tfh (cTfh) responses to R21/Matrix-M^®^. Cryopreserved PBMCs from Day 0 ((D0); baseline) and V3+7 (7 days post-third vaccination (V3)) were stained *ex vivo* and analysed by flow cytometry. Memory CD4+, CXCR5+PD1+ cTfh cells, and cTfh1/2/17 cells were defined as shown in Supplementary Figure 2. Frequencies were compared between **(A)** memory CD4+ T within total T cells at D0 and V3+7 within groups (**B**) cTfh (CXCR5+ PD1+) within total memory CD4+ T cells at D0 and V3+7 within groups, (**C**) memory CD4+ T cells between adults [Group 1A/B] and infants [Group 3B/D] at V3+28, and (**D**) cTfh cells between adults [Group 1A/B] and infants [Group 3B/D] at V3+28. (**E**) cTfh (CXCR5+ PD1+) phenotypes at D0 and V3+7 between adults [Group 1A/B] and infants [Group 3B/D]. Post-vaccination responses were compared within groups by Mann-Whitney test (**A-B**) or between groups (**C-E**). Sample sizes for all assays were based on sample availability; each point represents a single sample. Bars and error bars represent medians and interquartile ranges, respectively. Significant *p* values are annotated on graphs; *p* < 0.05 was considered significant.

In terms of differences between adults and infants, adult vaccinees had significantly higher frequencies of memory CD4+ T cells pre- and post-vaccination as compared to infant vaccinees (Figure 2C; D0 medians adults 1A/B 42% vs infants 3B/D 14.4% p = 0.001, and V3+7 medians adults 1A/B 42% vs infants 3B/D median 17.8% p = 0.0001). Within this CD4+ memory population, we observed no difference in frequencies of CXCR5+PD1+ cTfh cells in infants compared to adults (Figure 2D). There were also no significant differences in the frequencies of memory CD4+ T cells or CXCR5+PD1+ cTfh cells between the infant groups that received different R21 or Matrix-M^®^ doses (data not shown). However, we did observe higher frequencies of cTfh1 and lower frequencies of cTfh17 in infants as compared to adults within the CXRC5+PD1+ cTfh population, suggesting some differences in the cTfh cell repertoire (Figure 2E; Tfh1 medians adults 1A/B 11.25 % vs infants 3B/D 27.50% p = 0.0021, and Tfh17 medians adults 1A/B 19.70% vs infants 3B/D median 7.91% p = 0.0001). No correlation was observed between the frequencies of CXCR5+PD1+ cTfh cells at V3+7 and serum antibody at V3+28 or late time point (∼2 years post-vaccination; data not shown).

### Higher post-vaccination frequencies of RH5-specific cTfh cells were observed in children with higher previous exposure to malaria

In addition to our analyses of the licensed malaria vaccine R21/Matrix-M^®^, we were also interested in the vaccine antigen-specific cTfh and B cell responses to the leading blood-stage malaria vaccine candidate, RH5.1/Matrix-M ^®^. The groups from NCT04318002 included in this analysis are summarised in Table 1. In brief, this comprises Group 2A (children; 10µg RH5.1/ 50µg Matrix-M^®^ at 0-1-2 months), Group 2B (children; 10µg RH5.1/ 50µg Matrix-M^®^ at 0-1-6 months), Group 2C (children; 50µg RH5.1/ 50µg Matrix-M^®^ at 0-1 months and 10µg RH5.1/ 50µg Matrix-M^®^ at 6 months), and Group 2D (“high exposure” children; 50µg RH5.1/ 50µg Matrix-M^®^ at 0-1 months and 10µg RH5.1/ 50µg Matrix-M^®^ at 6 months).

Using cryopreserved samples from the RH5.1/Matrix-M^®^ trial, we were able to employ an AIM assay to report on RH5-specific cTfh frequencies. We first evaluated the baseline (Day 0; D0) and post-vaccination cTfh cell responses 7 days after the final vaccination in the primary three-dose series (V3+7) within total memory CD4+ T cells and total memory cTfh cells (as defined in Supplementary Figures 2-4). RH5-specific responses were detected within total memory CD4+ T cells in all groups (Figure 3A) and within total memory cTfh cells in all delayed regimen groups (Figure 3B). Trends were very similar when reporting frequencies of RH5-specific memory cTfh cells within CXCR5+PD1+ cells (Figure 3C). However, there were no significant bulk changes in the frequencies of total CXCR5+PD1+ cTfh cells within the memory CD4+ T cell population post-vaccination, except in the high malaria-exposed group (Supplementary Figure 8). Within cTfh1 (Figure 3D), cTfh2 (Figure 3E), and cTfh17 (Figure 3F) subsets (as defined in Supplementary Figures 2), RH5-specific responses were only detected consistently across all groups within the cTfh2 population (Figures 3E).

**Figure 3.**
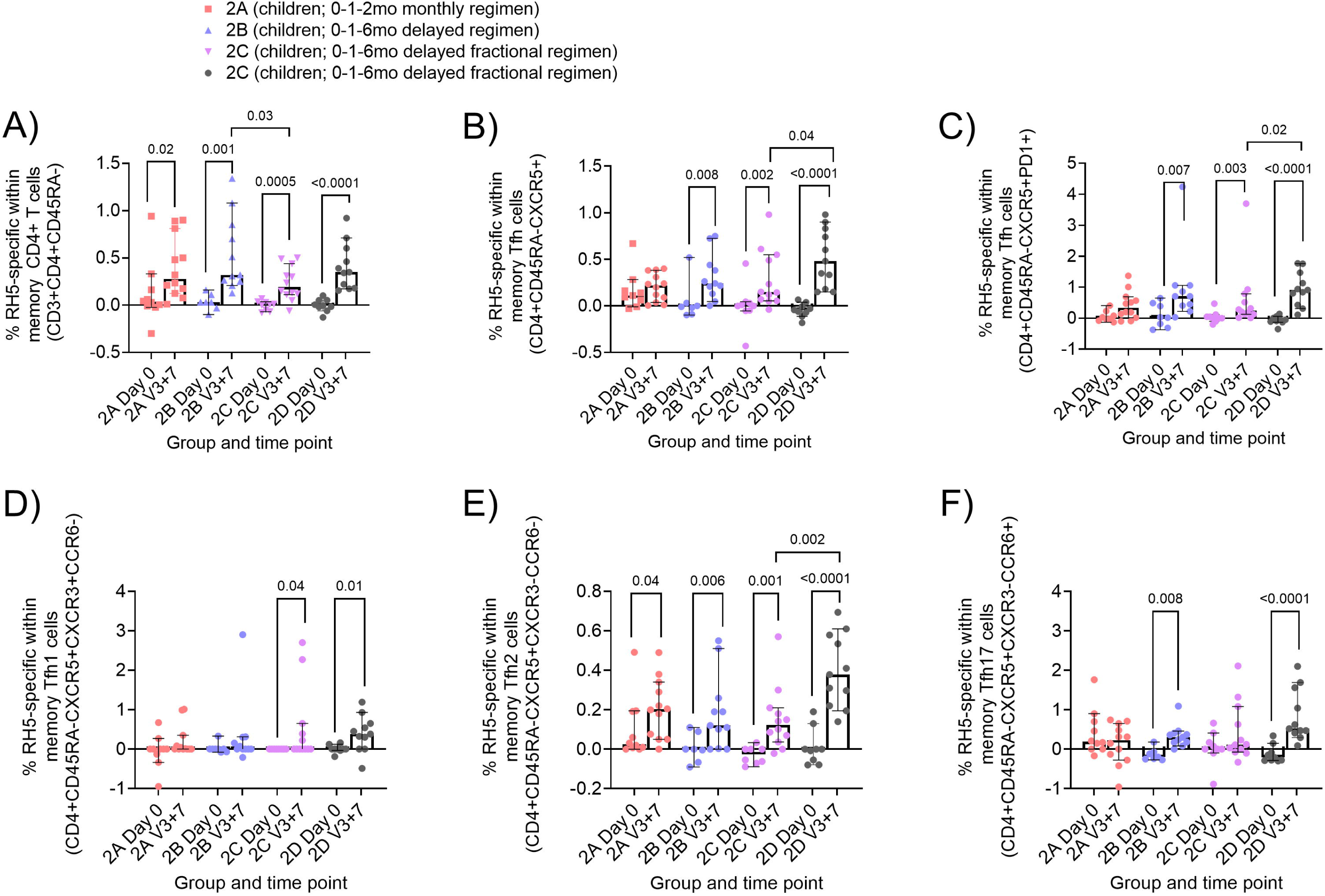
RH5-specific memory CD4+ and circulating Tfh (cTfh) responses to RH5.1/Matrix-M^®^. Cryopreserved PBMCs from Day 0 ((D0); baseline) and V3+7 (7 days post-third vaccination (V3)) were stimulated with medium alone or a RH5 peptide pool for 24h, then stained and analysed, identifying RH5-specific cells as those co-expressing CD25 with OX40 and/or CD69 following stimulation. RH5-specific memory T cells were identified as shown in Supplementary Figures 2-4). Frequencies were compared between (**A**) RH5-specific CD4+ T cells at D0 and V3+7 within groups, (**B**) RH5-specific cell responses within the total memory CXCR5+ cTfh at D0 and V3+7 within groups, (**C**) RH5-specific cells responses within total memory CXCR5+PD1+ cTfh at D0 and V3+7 within groups, (**D-F**) RH5-specific cTfh1, cTfh2 and cTfh17 at D0 and V3+7 within groups. The Mann-Whitney test was used for comparison between Day 0 (D0) and V3+7 (7 days post third vaccination), and between two groups. Sample sizes for all assays were based on sample availability; each point represents a single sample. Bars and error bars represent medians and interquartile ranges, respectively. Significant *p* values are annotated on graphs; p < 0.05 was considered significant. Note that negative data points are expected given that background responses to medium alone are subtracted from RH5-specific responses in matched samples in the AIM assay as described in Methods and published previously (Nielsen *et al*., 2021).

With respect to differences between groups, the comparisons we were interested in within the children vaccinees in the RH5.1/Matrix-M^®^ trial were Group 2A versus 2B (monthly versus delayed regimen), Group 2B versus 2C (delayed versus delayed fractional regimen), and Group 2C versus 2D (low versus high prior exposure to malaria; same delayed fractional regimen). Interestingly, within this RH5-specific cTfh cell data set, it was only in comparison between the different malaria pre-exposure groups that we observed differences in the magnitude of the RH5-specific cTfh cell response. Specifically, we observed higher frequencies of RH5-specific cells within the total memory CXCR5+ cTfh (Figure 3B V3+7 median 2D 0.48% vs 2C 0.15% p = 0.04), total memory CXCR5+PD1+ cTfh (Figure 3F V3+7 median 2D 0.88% vs 2C 0.19% p = 0.004), and memory cTfh2 (Figure 3D V3+7 median 2D 0.38% vs 2C 0.12% p = 0.001) populations in the “high exposure” Group 2D as compared to “low exposure” Group 2C who received the same delayed fractional regimen. Additionally, RH5-specific responses detected within total memory CD4+ T cells in the delayed regimen group (2B) were significantly higher than the delayed fractional regimen group (2C) post vaccination (Figure 3A V3+7 median 2B 0.32% vs 2C 0.19%, p = 0.03). In contrast to the *ex vivo* cTfh assay described for R21/Matrix-M^®^, no significant correlations were observed between frequencies of RH5-specific cTfh cells and anti-RH5.1 serum IgG at V3+28 (data not shown).

### Delayed booster vaccination drives higher RH5-specific memory IgG+ B cells than monthly or delayed fractional booster regimens

Within the RH5.1/Matrix-M^®^ trial (NCT04318002), RH5 probes were used to identify circulating RH5-specific IgG+, IgM+ and IgA+ memory B cells (defined as shown in Figure 4A and Supplementary Figure 1) in PBMC samples at baseline (Day 0; D0) and 28 days after the final vaccination in the primary three-dose series (V3+28). Significant RH5-specific total, resting, or activated memory IgG+ B cell responses were observed at V3+28 in all groups (total memory IgG+ Figure 4B; Supplementary Figure 9A and B). In contrast to the NANP analyses in R21/Matrix-M^®^ trial (Supplementary Figure 5), but consistent with previously published adult analyses from the same trial (Barrett *et al*., 2024), no significant IgM+ or IgA+ responses were detected post vaccination (Supplementary Figure 9C and D). However, at baseline, adults had a background signal with the RH5 probes in the IgM+ and IgA+ memory B cell subsets.

**Figure 4.**
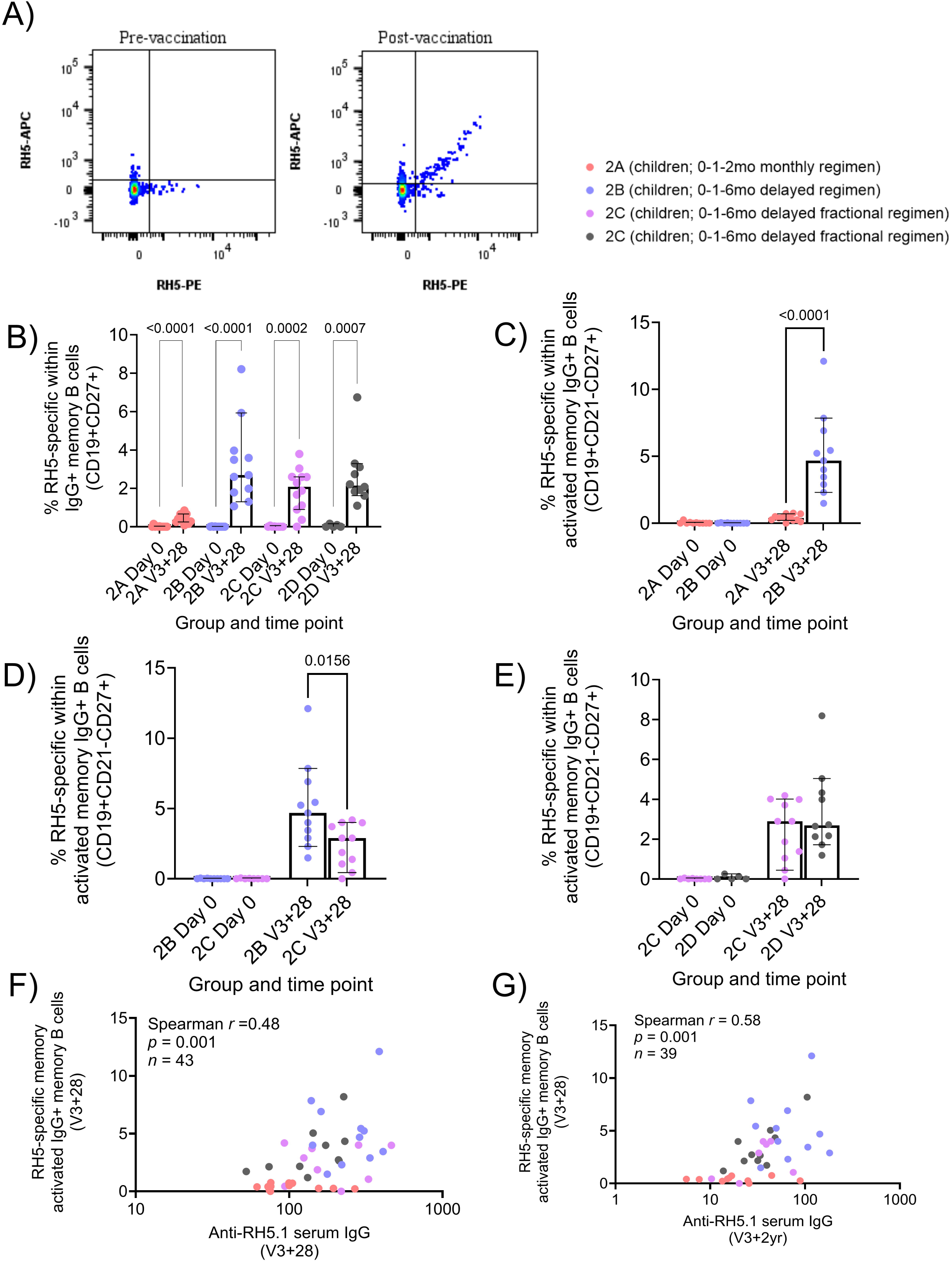
RH5-specific memory IgG+ B cells to RH5.1/Matrix-M^®^. Cryopreserved PBMCs from Day 0 (D0) and V3+28 (28 days post-third vaccination (V3)) were stained *ex vivo* and analysed by flow cytometry. RH5-specific memory B cell phenotypes were defined as shown in Figure (**A**) and Supplementary Figure 1. Frequencies were compared between (**B**) RH5-specific total IgG+ memory B cells at D0 and V3+28 within groups, (**C**) RH5-specific activated IgG+ memory B cells between “monthly” (Group 2A) vs “delayed” (Group 2B) vaccination regimen, (**D**) RH5-specific total IgG+ memory B cells between delayed (Group 2B) vs delayed fractional dose regimen (Group 2C), (**E**) RH5-specific total IgG+ memory B cells between low malaria pre-exposure (Group 2C) vs high malaria pre-exposure (Group 2D). Post-vaccination responses were compared within groups by the Mann-Whitney test (**B**) or between groups (**C-E**). A Spearman correlation analysis was performed between RH5-specific activated IgG+ mBC and anti-RH5.1 serum IgG at V3+28 and V3+2yr, respectively (**F-G**). Sample sizes for all assays were based on sample availability; each point represents a single sample. Bars and error bars represent medians and interquartile ranges, respectively. Significant p values are annotated on graphs; p < 0.05 was considered significant.

The delayed booster regimen induced substantially higher frequencies of RH5-specific B cells as compared to the monthly booster regimen across total, activated, and resting memory IgG+ populations (activated memory, Figure 4C; V3+28 medians: 0.40% Group 2A monthly, 4.68% Group 2B delayed, p = < 0.0001; Supplementary Figure 9E and F). Within activated memory IgG+ B cells, delayed boosting also appeared to induce a superior response than delayed fractional boosting (V3+28 median: 2.90% Group 2C delayed fractional, p = 0.0156; Figure 4D). There was no impact of high malaria pre-exposure on responses by any of the B cell populations measured (activated memory, Figure 4E; other data not shown). With respect to the relationship to serum antibody, RH5-specific responses within both total and activated memory IgG+ B cells correlated with matched time point serum anti-RH5.1 IgG (activated memory, Figure 4F, r = 0.48, p = 0.001; Supplementary Figure 9G). Unlike with NANP responses described above, these RH5-specific B cell responses continued to correlate strongly with serum IgG out to V3+2yr (Figure 4G, r = 0.58, p = 0.001, Supplementary Figure 9H).

### RH5.1/AS01 induced more durable antibodies in UK adults compared to RH5.1/Matrix-M^®^ in Tanzanian adults

Given these data on the strength of the RH5-specific B cell and serum antibody response in the context of delayed booster dosing of RH5.1/Matrix-M^®^, we contextualised these findings with previously published data on RH5.1. The anti-RH5.1 serum IgG kinetics from baseline to V3+2yr were replotted for both RH5.1/Matrix-M^®^ Tanzanian children vaccinees (Figure 5A; (Silk *et al*., 2024)) that have been the focus of the RH5 component of this study, as well as the Tanzanian RH5.1/Matrix-M^®^ adult vaccinees from the same trial, alongside RH5.1/AS01_B_ adult vaccinees from an earlier UK clinical trial (Figure 5B; NCT04318002 /(Minassian *et al*., 2021)).

**Figure 5.**
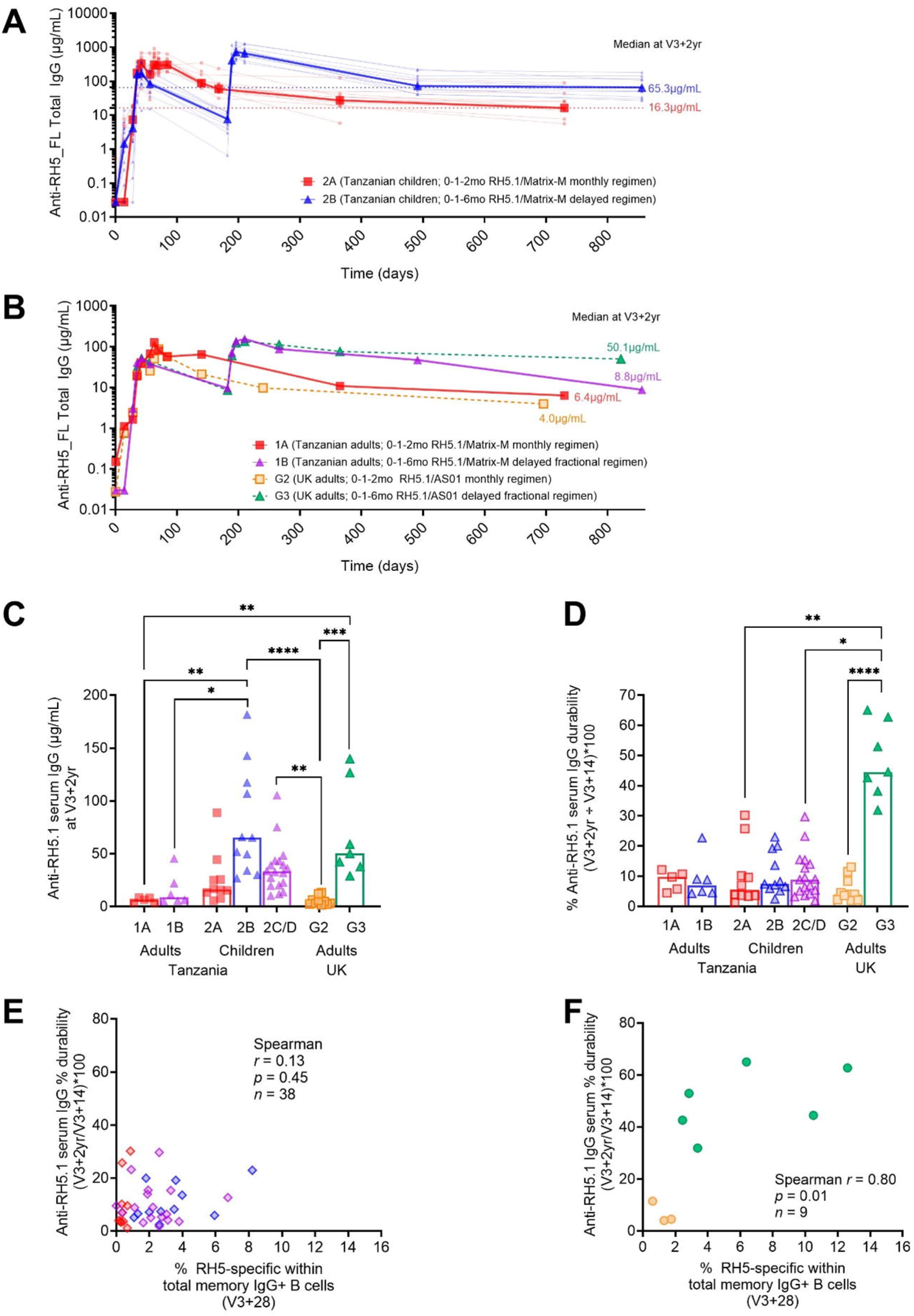
Comparison of Tanzanian RH5.1/Matrix-M^®^ serum IgG and B cell responses to previous RH5.1/AS01_B_ UK clinical trial. Full anti-RH5.1 serum IgG kinetics from baseline to 2 years post-third vaccination (FV+2yr) was measured by ELISA in both the RH5.1/Matrix-M^®^ (NCT04318002) (Silk *et al*., 2024) and RH5.1/AS01_B_ (NCT02927145) clinical trials (Minassian *et al*., 2021). Here we draw attention to the kinetics between “monthly” (2A) and “delayed” (2B) groups in Tanzanian children receiving RH5.1/Matrix-M^®^ (**A**) and between “monthly” (1A), “delayed fractional” (1B) Tanzanian adults receiving RH5.1/Matrix-M^®^, and “monthly” (G2) or “delayed fractional” (G3) UK adults receiving RH5.1/AS01_B_ (**B**). Anti-RH5.1 serum IgG at V3+2yr was compared between groups in both trials (**C**), as well as percentage durability of anti-RH5.1 serum IgG (a read-out of serum IgG durability/ persistence/ decay) between V3+14 and V3+2yrs (**D**). A Kruskal Wallis test was used to compare the mean rank of each group with the mean rank of every other group with Dunn’s correction for multiple comparisons (**C-D**). Spearman correlation analyses were performed between percentage durability of anti-RH5.1 serum IgG with RH5-specific total memory IgG+ B cells (V3+28) in children receiving RH5.1/Matrix-M^®^ (NCT04318002; **E**) and adults receiving RH5.1/AS01_B_ (NCT02927145; **F**). In all graphs, red = monthly regimens with Matrix-M^®^, blue = delayed regimens with Matrix-M^®^, purple = delayed fractional regimens with Matrix-M^®^, orange = monthly regimens with AS01_B_, and green = delayed fractional regimens with AS01_B_. Sample sizes for all assays were based on sample availability; each point represents a single sample with the exception of (**A**) where thick lines and large points represent group medians and (**B**) where all lines and points represent group medians. Bars and error bars represent medians and interquartile ranges, respectively (**C-D**). Significant p values are annotated on graphs; p < 0.05 was considered significant.

While delayed boosting with Matrix-M^®^ in Tanzanian children or delayed fractional boosting with AS01_B_ in UK adults results in higher anti-RH5.1 serum IgG at V3+2yr as compared to monthly regimens, we did not see the same effect in Tanzanian adults receiving delayed fractional boosting with Matrix-M^®^. Only the Tanzanian children who received the delayed regimen (2B) with Matrix-M® and the UK adults who received the delayed fractional regimen (G3) with AS01_B_ maintained median anti-RH5.1 IgG concentrations above 50 µg/mL at V3+2yr (Figure 5C). However, the mechanism behind these high V3+2yr anti-RH5.1 titres appears to be different: in the delayed fractional dosing AS01_B_ UK adults, the serum IgG percentage durability was approximately four-fold higher than in any other regimen (median = 44.5% of peak anti-RH5.1 µg/mL still present at V3+2yr versus < 10% in all other groups; Figure 5D). Conversely, the high V3+2yr titres in Tanzanian children seemed linked to a more robust peak response in the first instance, not an improvement in serum antibody persistence. Consistent with this, both V3+28 B cell and serum IgG responses correlate with late time point V3+2yr serum antibody (Figure 4F) but not percentage durability (Figure 5E) in the Tanzanian children. In contrast, the V3+28 B cell response in UK adults with AS01_B_ correlates with both late time point V3+2yr serum antibody and percentage durability (Figure 5F).

There is perhaps a yet to be understood interaction between adjuvant, age, antigen-specific B cell activation and serum antibody persistence. However, we also note that these are small sample sizes and that the median timing of the V3+2yr visit was earlier in the G2 and G3 RH5.1/AS01_B_ UK groups as compared to the RH5.1/Matrix-M^®^ Tanzanian groups due to differences in operational use of approved sampling windows during the respective clinical trials (NCT04318002 G2/G3 median = V3+510 days [range 386-826]; NCT04318002 median = V3+681 days [range 675-698]). Further work is therefore needed to validate these findings, ideally with larger sample sizes and – for the flow cytometry – head-to-head comparisons within the same assays.

## Discussion

In this study, we investigated the impact of multiple parameters on the induction of vaccine-specific B and cTfh cell responses to R21/Matrix-M^®^ and candidate RH5.1/Matrix-M^®^ malaria vaccines. Using PBMC samples from the R21/Matrix-M^®^ trial in Kenya (NCT03580824) and the RH5.1/Matrix-M^®^ trial in Tanzania (NCT04318002), we were able to identify differences in immunogenicity by timing of booster vaccinations (RH5.1), prior exposure to malaria (RH5.1), adjuvant dose (R21), and age (R21).

The impact of delayed booster dosing on B cell immunogenicity and resultant serum antibody responses to RH5.1 remains an intriguing topic. We have previously published enhanced B cell immunogenicity with delayed fractional booster dosing in Tanzanian adults receiving RH5.1/Matrix-M^®^ (Barrett *et al*., 2024; Silk *et al*., 2024), and in UK adults receiving RH5.1/AS01_B_ (Minassian *et al*., 2021; Nielsen *et al*., 2023). Here, we present the first data on the effect of RH5.1 delayed boosting on B cell responses in children living in a malaria-endemic setting. We observed an increased IgG+ memory B cell response with delayed booster dosing compared to monthly dosing regimen or delayed fractional dosing, which correlated with both matched peak (V3+28) and late (V3+2yr) time point serum antibody. Additionally, a delayed booster induced higher RH5-specific IgG+ memory B cells than a delayed fractional dose in children. This is the first report on the head-to-head comparison of delayed vs delayed fractional dose impact on RH5-specific memory B cells and is highly supportive of further clinical development of the delayed boosting regimen. The mechanism by which delayed booster dosing induces higher B cell responses is not fully understood, but is likely related to enhanced germinal centre responses, possibly driven by increased competition for antigen following a delayed final booster (Asante *et al*., 2011; Shlomchik and Weisel, 2012; Regules *et al*., 2016; McCall, Yap and Bousema, 2020), or waning negative feedback from circulating serum antibody and ongoing germinal centres induced by the second vaccination (Nielsen *et al*., 2021; Barrett *et al*., 2024). Ongoing studies using fine needle aspiration of draining lymph nodes aim to directly examine these mechanisms (NCT05978037; NCT06320535).

The analysis of NANP-specific memory B cell responses post-vaccination with R21/Matrix-M^®^ revealed interesting heterogeneity, with higher responses in infants compared to adults in Kenya. This could be associated with the effect of previous malaria exposure since this cohort was recruited from a region of moderate malaria endemicity (Junju, Kenya) (Bejon *et al*., 2010). Previously, the frequency of total B cells was reported to be 1.4-fold lower in individuals with previous malaria exposure compared to those with primary infection (Sundling *et al*., 2019). However, immunomodulatory effects of chronic malaria exposure are well documented (Illingworth *et al*., 2013; Obeng-Adjei *et al*., 2015; Bediako *et al*., 2019). In addition, there was a significant increase in responses above baselines in all isotypes (IgG+, IgA+, and IgM+ B cells) in contrast to RH5. This could be an indication of a difference between RH5.1/Matrix-M^®^ and R21/Matrix-M^®^ vaccine responses, for example, related to soluble protein vs nanoparticle platforms, respectively. Our findings align with a recent report showing increased C-terminus and full-length R21-specific memory B cells after R21 vaccination across IgG, IgM and IgA isotypes in malaria-exposed Kenyan adults, as measured by Fluorospot (Kibwana *et al*., 2025). Analyses of NANP-specific memory B cells were not included in this prior study despite the central role of anti-NANP antibodies in R21/Matrix-M® vaccine development and licensure (Datoo *et al*., 2024).

In the R21/Matrix-M^®^ cohort, vaccination with a higher dose of Matrix-M^®^ was observed to drive more activated B cells in infant vaccinees. Previously, a higher Matrix-M^®^ dose (50µg) has been associated with increased clinical efficacy, and higher magnitude of NANP IgG antibodies in Burkinabe children (Datoo *et al*., 2021) and higher complement fixing capacity and sporozoite inhibitory capabilities compared to the lower Matrix-M^®^ dose (25µg) in Kenyan children (Bundi et al., 2025). We hypothesise that the higher Matrix-M^®^ dose, and not the vaccine antigen dose, improved germinal centre activity by enhancing vaccine antigen uptake and presentation (Reimer *et al*., 2012), resulting in higher frequencies of total and activated NANP-specific IgG+ memory B cells in circulation. These data emphasise the importance of optimising the adjuvant dose for vaccine immunogenicity (Facciolà *et al*., 2022). While all RH5.1/Matrix-M^®^ vaccinees received 50 µg Matrix-M^®^, precluding any analysis of the impact of adjuvant dose, these R21/Matrix-M^®^ data indicate that decreasing the Matrix-M^®^ adjuvant dose would likely have a negative impact on RH5.1 immunogenicity.

The goal of this work was not to directly compare R21 and RH5.1 vaccine responses, but differences in the magnitude of antigen-specific memory B cell responses at V3+28 are noteworthy. In infants receiving R21/Matrix-M® (NCT03580824), the highest median NANP-specific activated IgG+ memory B cell response (Group 3B/D; 0.56%) was eight-fold lower than that observed in RH5.1/Matrix-M® Group 2B delayed regimen vaccinees (NCT04318002; 4.68%), though comparable to the lowest monthly regimen (Group 2A; 0.40%). As delayed dosing has not been evaluated for R21, it remains unclear whether a delayed booster would elicit responses similar to those seen with RH5.1. Data from different dosing regimens with the licensed pre-erythrocytic vaccine RTS,S/AS01_B_ (also a CSP-based nanoparticle) are mixed, suggesting delayed boosting with an arrayed repeat epitope, such as a CSP-based nanoparticle vaccine, may behave differently from delayed boosting with a protein antigen, such as RH5.1. For example, while delayed fractional dosing with RTS,S/AS01_B_ has been associated with higher frequencies of memory B cells compared to a monthly schedule (Pallikkuth *et al*., 2020), no delayed boosting regimens have yet been linked to higher efficacy in endemic settings (Samuels *et al*., 2022). To note, in delayed fractional RTS,S/AS01_B_ groups both the RTS,S antigen and AS01 adjuvant were fractionated, whereas in our RH5.1 trials, only the antigen is fractionated while the adjuvant dose (AS01 or Matrix-M®) is held constant.

Similarly, we were not intending to directly compare the magnitude of vaccine-specific B cell responses to historical B cell flow cytometry datasets from UK adults with RH5.1/AS01_B_ (NCT02927145) or Tanzanian adults with RH5.1/Matrix-M^®^ (NCT04318002), given that the assays were run years apart with slightly different protocols and cytometer settings. However, trends remain consistent with higher frequencies of RH5-specific IgG+ memory B cells observed with delayed booster regimens in both trials (Barrett et al., 2024; Nielsen et al., 2023).

Given the critical role of Tfh in supporting the GC B cell response, there is significant interest in efficiently eliciting Tfh responses to vaccine antigens (Yu *et al*., 2022). Following R21/Matrix-M® vaccination, we observed increased frequencies of bulk circulating Tfh (cTfh) cells in adults, consistent with findings in Kenyan adults showing elevated CXCR5+PD1+ cTfh among memory CD4+ T cells two weeks post-vaccination (Kibwana *et al*., 2025). No significant differences were observed in infants or between adults and infants at baseline or post-vaccination. However, within CXCR5+PD1+ cTfh, infants displayed higher proportions of cTfh1 and lower cTfh17 subsets compared to adults post-vaccination. This aligns with previous observations that cTfh1 phenotypes predominate in children, while cTfh2 phenotypes are more frequent in adults (Chan *et al*., 2022).

In the RH5.1/Matrix-M^®^ trials, the vaccination dose or timing of final (third) vaccine booster did not impact the RH5-specific cTfh cell response. Similarly, in a prior RH5.1/AS01_B_ trial (NCT02927145) in malaria-naïve UK adults, no significant difference was found between delayed and monthly dosing schedules (Minassian *et al*., 2021). This consistent with observations from SARS-CoV-2 mRNA vaccine, where delayed boosting had no effect on the CD4+ T cells (Nicolas *et al*., 2023). In contrast, delayed boosting enhanced CSP-specific cTfh frequencies in malaria naïve adults vaccinated with RTS,S/AS01_B_ (0-1-7mo versus 0-1-2mo)(Pallikkuth *et al*., 2020). Surprisingly, RH5-specific cTfh and cTfh2 frequencies were higher in malaria pre-exposed (“high exposure”) than in “low exposure” children. Although chronic malaria exposure has been associated with impaired immune systems (Illingworth *et al*., 2013; Ubillos *et al*., 2017), in this cohort the children were young (5-17 months) and thus may not have sufficiently prolonged/ chronic exposure required to impair the immune system, especially given that exposure levels in Tanzania are overall low in the wider African context (see discussion and methods in the original trial publication (Silk *et al*., 2024). The higher RH5-specific cTfh and cTfh2 responses seen among “high exposure” children could possibly be associated with low existing memory boosted by the vaccine, but given that RH5 responses to natural exposure are very low, this seems unlikely (Silk *et al*., 2023, 2024; Natama *et al*., 2025). Consistent with this, no difference was detected in RH5-specific B cell or serum IgG responses by exposure level, suggesting no impairment to the humoral response. Similar analyses in higher endemicity settings may be of interest.

One main challenge facing any malaria vaccine is maintenance of vaccine-induced immunity (RTS, 2015; Olotu *et al*., 2016; Minassian *et al*., 2021; Datoo *et al*., 2022). Published data has shown that RH5.1/AS01_B_ vaccination of UK adults (Minassian *et al*., 2021) resulted in higher anti-RH5.1 serum IgG at V3+2yr compared to Tanzanian adults vaccinated with RH5.1/Matrix-M^®^ (Silk *et al*., 2024). While demographic (malaria naïve vs exposed) and timing of V3+2yr sampling differences may contribute to this, adjuvant effects on antibody durability are also possible. For example, with RH5.1/Matrix-M^®^, peak vaccine-specific B cell and serum antibody correlate with late time point antibody (i.e., better responses at peak indicative of better maintained serum IgG after 2 years), but not with percentage serum antibody durability (i.e., rate of decay of serum antibody). In contrast, RH5.1/AS01_B_ data shows correlations both between peak time point B cells and also serum antibody maintenance. This suggests further work is needed to understand how adjuvants interact with dosing regimen to affect the durability of humoral immunogenicity. Extending our pilot bone marrow sampling work to look at long-lived plasma cell induction with different adjuvant/ dosing regimen combination would be useful (Barrett *et al*., 2024) as would draining lymph node sampling to assess germinal centre responses and systems vaccinology approaches to better understand early biomarkers of vaccine-induced antibody responses and whether these differ between vaccine platforms (Hagan *et al*., 2022; Cortese *et al*., 2025). Serology and B cell data from monthly, delayed, and delayed fractional regimens with RH5.1/ Matrix-M^®^ in ongoing Phase I trials (NCT05385471, NCT06141057) in healthy malaria-naïve UK adults will also fill in important gaps needed for improving our understanding of this potential adjuvant/ dosing regimen interaction. The first multi-stage malaria vaccine clinical trials – testing RH5.1/ Matrix-M^®^ and R21/Matrix-M^®^ – are currently ongoing and include both vaccination regimens where RH5.1 and R21 vaccines are administered at different visits (NCT05790889), and also regimens where both vaccine antigens are administered at the same visit and “share” the Matrix-M^®^ dose (NCT05357560). To note, if the beneficial impact of delayed booster dosing is ultimately found to be adjuvant-dependent (AS01 or similar), this may present complications for co-formulation/ co-administration of blood-stage vaccines with pre-erythrocytic vaccines that utilise different adjuvants, e.g., R21/Matrix-M^®^ or sporozoite candidates.

Our study had a few limitations. First, we used samples from Phase 1b malaria vaccine trials, which involved small numbers of participants (n <20 per group) (Sang *et al*., 2023; Silk *et al*., 2023), and further reduced by limited PBMC vial availability. Therefore, statistical power for between-group comparisons was low, and analyses could not account for potential confounders such as sex. Replication with larger datasets will be essential to validate our findings. PBMC availability also affected the time points used in the B cell analyses; as V3+7 was used for the cTfh cell assays we were unable to also analyse B cell responses at this time point, precluding our capacity to report short-lived plasma cell responses which peak at this time point, but are transient (Barrett *et al*., 2024). Previous pilot studies have explored the option of splitting vials between assays since the B cell assay described here uses negatively enriched B cells (i.e., the B cell-depleted PBMC fraction could be used for a T cell assay), but preliminary data suggested this might negatively affect background in the AIM assay. While it would have been of considerable interest to assess the impact of various parameters, such as timing of booster vaccinations, prior malaria exposure, adjuvant dose, and age, on immune responses to both R21/Matrix-M^®^ and RH5.1/Matrix-M^®^, such analyses were limited by the constraints of the original study design.

To conclude, this study provides the first direct analysis of vaccine antigen-specific B cell responses in the target population for both the licensed pre-erythrocytic vaccine R21/Matrix-M® and the leading blood-stage candidate RH5.1/Matrix-M®. Taken together, this work indicates that age, adjuvant dose, vaccine dose, and timing of final booster vaccination impact the magnitude of the antigen-specific B cell response. We also previously observed this in antibody responses (Silk *et al*., 2024; Bundi *et al*., 2025). The cTfh cell data were less clear and overall suggest that these modifiable vaccine parameters have a greater influence on B cell rather than T cell immunogenicity. Comparison to historical RH5.1/AS01_B_ data further implies that adjuvant choice (Matrix-M® vs AS01) may influence the relationship between peak B cell responses and long-term antibody persistence, though current RH5.1/AS01_B_ data remain limited to a small malaria-naïve adult cohort(no children or infants).

Regardless, there is clear scope for a discussion in the malaria vaccine field regarding the importance of declining serum antibody concentrations over time: if two regimens induce equivalent antibody concentrations after 2 years is it biologically relevant if decay kinetics are different? Should vaccine development programmes seek to optimise peak responses, late time point concentrations, or limit the rate of decline between the two? Answering such questions will likely require modelling data from Phase 2b efficacy trials where antibody concentrations can be evaluated alongside clinical episodes. Likewise, further cellular analyses in larger, independent clinical trials will be important to help us further understand the impact of these modifiable vaccine parameters to guide multi-stage malaria vaccine optimisation, as well as the identification of putative early B cell biomarkers of durability of humoral immunity and protection.

## Data Availability

All data produced in the present study are available upon reasonable request to the authors.

## Acknowledgments

We thank the clinical trial participants and wider clinical trial teams for participating in and running the clinical trials essential for this study. For NCT04318002, we especially thank Fay Nugent, Jee-Sun Cho, Rachel Roberts, Saumu Ahmed, Florence Milando, and Neema Balige. We also thank Jenny Reimer and Cecilia Carnot at Novavax for providing the Matrix-M^®^ adjuvant. For NCT03580824 we thank Emmaloise Gathuri, Omar Ngoto and Janet Musembi. We thank Robert Hedley and Vasiliki Tsioligka for providing technical assistance in the analysis of the NCT04318002 samples at the Don Mason Facility of Flow Cytometry, Sir William Dunn School of Pathology, University of Oxford. We thank Andrew Worth from the Jenner Institute for offering technical support in the analysis of NCT04318002 and NCT03580824 flow cytometry.

## References

1. Asante, K.P. et al. (2011) ‘Safety and efficacy of the RTS,S/AS01E candidate malaria vaccine given with expanded-programme-on-immunisation vaccines: 19 month follow-up of a randomised, open-label, phase 2 trial’, The Lancet Infectious Diseases, 11(10). Available at: 10.1016/S1473-3099(11)70100-1.

2. Barrett, J.R. et al. (2024) ‘Analyses of human vaccine-specific circulating and bone marrow-resident B cell populations reveal benefit of delayed vaccine booster dosing with blood-stage malaria antigens’, Frontiers in Immunology, 14. Available at: 10.3389/fimmu.2023.1193079.

3. Bediako, Y. et al. (2019) ‘Repeated clinical malaria episodes are associated with modification of the immune system in children’, BMC Medicine, 17(1). Available at: 10.1186/s12916-019-1292-y.

4. Bejon, P. et al. (2010) ‘Stable and unstable malaria hotspots in longitudinal cohort studies in Kenya.’, PLoS medicine, 7(7), p. e1000304. Available at: 10.1371/journal.pmed.1000304.

5. Bowyer, G. et al. (2018) ‘CXCR3+ T follicular helper cells induced by co-administration of RTS, S/AS01B and viral-vectored vaccines are associated with reduced immunogenicity and efficacy against malaria’, Frontiers in Immunology, 9(JUL), pp. 1–15. Available at: 10.3389/fimmu.2018.01660.

6. Brenna, E. et al. (2020) ‘CD4+ T Follicular Helper Cells in Human Tonsils and Blood Are Clonally Convergent but Divergent from Non-Tfh CD4+ Cells’, Cell Reports, 30(1). Available at: 10.1016/j.celrep.2019.12.016.

7. Bundi, C. et al. (2025) ‘Vaccine-induced responses to R21/Matrix-M – an analysis of samples from a phase 1b age de-escalation, dose-escalation trial’, Frontiers in Immunology, 16. Available at: 10.3389/fimmu.2025.1620366.

8. Chan, J.A. et al. (2022) ‘Age-dependent changes in circulating Tfh cells influence development of functional malaria antibodies in children’, Nature Communications, 13(1). Available at: 10.1038/s41467-022-31880-6.

9. Collins, K.A. et al. (2017) ‘Enhancing protective immunity to malaria with a highly immunogenic virus-like particle vaccine’, Scientific Reports, 7(April), pp. 1–15. Available at: 10.1038/srep46621.

10. Cortese, M. et al. (2025) ‘System vaccinology analysis of predictors and mechanisms of antibody response durability to multiple vaccines in humans’, Nature Immunology, 26(1), pp. 116–130. Available at: 10.1038/s41590-024-02036-z.

11. Crosnier, C. et al. (2013) ‘A library of functional recombinant cell-surface and secreted p. Falciparum merozoite proteins’, Molecular and Cellular Proteomics, 12(12), pp. 3976–3986. Available at: 10.1074/mcp.O113.028357.

12. Crotty, S. (2014) ‘T follicular helper cell differentiation, function, and roles in disease.’, Immunity, 41(4), pp. 529–542. Available at: 10.1016/j.immuni.2014.10.004.

13. Datoo, M.S. et al. (2021) ‘Efficacy of a low-dose candidate malaria vaccine, R21 in adjuvant Matrix-M, with seasonal administration to children in Burkina Faso: a randomised controlled trial.’, *Lancet (London*, England), 397(10287), pp. 1809–1818. Available at: 10.1016/S0140-6736(21)00943-0.

14. Datoo, M.S. et al. (2022) ‘Efficacy and immunogenicity of R21/Matrix-M vaccine against clinical malaria after 2 years’ follow-up in children in Burkina Faso: a phase 1/2b randomised controlled trial.’, The Lancet. Infectious diseases, 0(0). Available at: 10.1016/S1473-3099(22)00442-X.

15. Datoo, M.S. et al. (2024) ‘Safety and efficacy of malaria vaccine candidate R21/Matrix-M in African children: a multicentre, double-blind, randomised, phase 3 trial’, The Lancet, 403(10426). Available at: 10.1016/S0140-6736(23)02511-4.

16. Dicko, A. et al. (2024) ‘Seasonal vaccination with RTS,S/AS01E vaccine with or without seasonal malaria chemoprevention in children up to the age of 5 years in Burkina Faso and Mali: a double-blind, randomised, controlled, phase 3 trial’, The Lancet Infectious Diseases, 24(1). Available at: 10.1016/S1473-3099(23)00368-7.

17. van Dorst, M.M.A.R. et al. (2024) ‘Immunological factors linked to geographical variation in vaccine responses’, Nature Reviews Immunology. Available at: 10.1038/s41577-023-00941-2.

18. Facciolà, A. et al. (2022) ‘An Overview of Vaccine Adjuvants: Current Evidence and Future Perspectives’, Vaccines. Available at: 10.3390/vaccines10050819.

19. Hagan, T. et al. (2022) ‘Transcriptional atlas of the human immune response to 13 vaccines reveals a common predictor of vaccine-induced antibody responses’, Nature Immunology, 23(12), pp. 1788–1798. Available at: 10.1038/s41590-022-01328-6.

20. Hill, D.L. et al. (2019) ‘The adjuvant GLA-SE promotes human Tfh cell expansion and emergence of public TCRβ clonotypes.’, The Journal of experimental medicine, 216(8), pp. 1857–1873. Available at: 10.1084/jem.20190301.

21. Hodgson, D. et al. (2025) ‘Memory B cell proliferation drives differences in neutralising responses between ChAdOx1 and BNT162b2 SARS-CoV-2 vaccines’, Frontiers in Immunology, Volume 16-2025. Available at: https://www.frontiersin.org/journals/immunology/articles/10.3389/fimmu.2025.1487066.

22. Huber, J.E. et al. (2020) ‘Dynamic changes in circulating T follicular helper cell composition predict neutralising antibody responses after yellow fever vaccination’, Clinical and Translational Immunology, 9(5). Available at: 10.1002/cti2.1129.

23. Illingworth, J. et al. (2013) ‘Chronic Exposure to Plasmodium falciparum Is Associated with Phenotypic Evidence of B and T Cell Exhaustion’, The Journal of Immunology [Preprint], (24). Available at: 10.4049/jimmunol.1202438.

24. Kibwana, E. et al. (2025) ‘R21/Matrix-M malaria vaccine drives diverse immune responses in pre-exposed adults: insights from a phase IIb controlled human malaria infection trial’, Frontiers in Immunology, 16. Available at: 10.3389/fimmu.2025.1620365.

25. Koutsakos, M. et al. (2018) ‘Circulating TFH cells, serological memory, and tissue compartmentalization shape human influenza-specific B cell immunity’, Science Translational Medicine, 10(428). Available at: 10.1126/scitranslmed.aan8405.

26. Laurens, M.B. (2020) ‘RTS,S/AS01 vaccine (Mosquirix^TM^): an overview’, Human Vaccines and Immunotherapeutics, 16(3). Available at: 10.1080/21645515.2019.1669415.

27. Locci, M. et al. (2013) ‘Human circulating PD-1+CXCR3-CXCR5+ memory Tfh cells are highly functional and correlate with broadly neutralizing HIV antibody responses.’, Immunity, 39(4), pp. 758–769. Available at: 10.1016/j.immuni.2013.08.031.

28. McCall, M.B.B., Yap, X.Z. and Bousema, T. (2020) ‘Optimizing RTS,S Vaccination Strategies: Give It Your Best Parting Shot’, The Journal of Infectious Diseases, 222(10), pp. 1581–1584. Available at: 10.1093/infdis/jiaa423.

29. Minassian, A.M. et al. (2021) ‘Reduced blood-stage malaria growth and immune correlates in humans following RH5 vaccination’, Med [Preprint]. Available at: 10.1016/j.medj.2021.03.014.

30. Mudd, P.A. et al. (2022) ‘SARS-CoV-2 mRNA vaccination elicits a robust and persistent T follicular helper cell response in humans’, Cell, 185(4). Available at: 10.1016/j.cell.2021.12.026.

31. Natama, H.M. et al. (2025) ‘Safety and efficacy of the blood-stage malaria vaccine RH5.1/Matrix-M in Burkina Faso: interim results of a double-blind, randomised, controlled, phase 2b trial in children’, The Lancet Infectious Diseases, 25(5), pp. 495–506. Available at: 10.1016/S1473-3099(24)00752-7.

32. Nicolas, A. et al. (2023) ‘An extended SARS-CoV-2 mRNA vaccine prime-boost interval enhances B cell immunity with limited impact on T cells’, iScience, 26(1). Available at: 10.1016/j.isci.2022.105904.

33. Nielsen, C.M. et al. (2021) ‘Protein/AS01B vaccination elicits stronger, more Th2-skewed antigen-specific human T follicular helper cell responses than heterologous viral vectors’, Cell Reports Medicine, 2(3), p. 100207. Available at: 10.1016/j.xcrm.2021.100207.

34. Nielsen, C.M. et al. (2023) ‘Delayed boosting improves human antigen-specific Ig and B cell responses to the RH5.1/AS01B malaria vaccine’, JCI Insight, 8(2). Available at: 10.1172/jci.insight.163859.

35. Obeng-Adjei, N. et al. (2015) ‘Circulating Th1-Cell-type Tfh Cells that Exhibit Impaired B Cell Help Are Preferentially Activated during Acute Malaria in Children’, Cell Reports, 13(2), pp. 425–439. Available at: 10.1016/j.celrep.2015.09.004.

36. Olotu, A. et al. (2016) ‘Seven-Year Efficacy of RTS,S/AS01 Malaria Vaccine among Young African Children.’, The New England journal of medicine, 374(26), pp. 2519–2529. Available at: 10.1056/NEJMoa1515257.

37. Pallikkuth, S. et al. (2020) ‘A delayed fractionated dose RTS,S AS01 vaccine regimen mediates protection via improved T follicular helper and B cell responses’, eLife, 9, p. e51889. Available at: 10.7554/eLife.51889.

38. Palm, A.K.E. and Henry, C. (2019) ‘Remembrance of Things Past: Long-Term B Cell Memory After Infection and Vaccination’, Frontiers in immunology. Available at: 10.3389/fimmu.2019.01787.

39. Payne, R.O. et al. (2017) ‘Human vaccination against RH5 induces neutralizing antimalarial antibodies that inhibit RH5 invasion complex interactions’, JCI Insight, 2(21), pp. 1–19. Available at: 10.1172/jci.insight.96381.

40. Qi, H. (2016) ‘T follicular helper cells in space-time’, Nature Reviews Immunology. Available at: 10.1038/nri.2016.94.

41. Regules, J.A. et al. (2016) ‘Fractional Third and Fourth Dose of RTS,S/AS01 Malaria Candidate Vaccine: A Phase 2a Controlled Human Malaria Parasite Infection and Immunogenicity Study.’, The Journal of infectious diseases, 214(5), pp. 762–771. Available at: 10.1093/infdis/jiw237.

42. Reimer, J.M. et al. (2012) ‘Matrix-m^TM^ adjuvant induces local recruitment, activation and maturation of central immune cells in absence of antigen’, PLoS ONE, 7(7). Available at: 10.1371/journal.pone.0041451.

43. RTS, S.C.T.P. (2015) ‘Efficacy and safety of RTS,S/AS01 malaria vaccine with or without a booster dose in infants and children in Africa: Final results of a phase 3, individually randomised, controlled trial’, The Lancet, 386(9988). Available at: 10.1016/S0140-6736(15)60721-8.

44. Samuels, A.M. et al. (2022) ‘Efficacy of RTS,S/AS01E malaria vaccine administered according to different full, fractional, and delayed third or early fourth dose regimens in children aged 5–17 months in Ghana and Kenya: an open-label, phase 2b, randomised controlled trial’, The Lancet Infectious Diseases, 22(9). Available at: 10.1016/S1473-3099(22)00273-0.

45. Sang, S. et al. (2023) ‘Safety and immunogenicity of varied doses of R21/Matrix-M^TM^ vaccine at three years follow-up: A phase 1b age de-escalation, dose-escalation trial in adults, children, and infants in Kilifi-Kenya’, Wellcome Open Research, 8. Available at: 10.12688/wellcomeopenres.19795.1.

46. Shchelkunova, G.A. and Shchelkunov, S.N. (2017) ‘40 Years without Smallpox’, Acta Naturae, 9(4). Available at: 10.32607/20758251-2017-9-4-4-12.

47. Shlomchik, M.J. and Weisel, F. (2012) ‘Germinal center selection and the development of memory B and plasma cells’, Immunological Reviews, 247(1). Available at: 10.1111/j.1600-065X.2012.01124.x.

48. Silk, S.E. et al. (2023) ‘Superior antibody immunogenicity of a viral-vectored RH5 blood-stage malaria vaccine in Tanzanian infants as compared to adults’, Med [Preprint]. Available at: 10.1016/j.medj.2023.07.003.

49. Silk, S.E. et al. (2024) ‘Blood-stage malaria vaccine candidate RH5.1/Matrix-M in healthy Tanzanian adults and children; an open-label, non-randomised, first-in-human, single-centre, phase 1b trial’, The Lancet Infectious Diseases [Preprint]. Available at: 10.1016/S1473-3099(24)00312-8.

50. De Silva, N.S. and Klein, U. (2015) ‘Dynamics of B cells in germinal centres.’, Nature reviews. Immunology, 15(3), pp. 137–148. Available at: 10.1038/nri3804.

51. Sundling, C. et al. (2019) ‘B cell profiling in malaria reveals expansion and remodeling of CD11c+ B cell subsets’, JCI Insight, 4(9). Available at: 10.1172/jci.insight.126492.

52. Ubillos, I. et al. (2017) ‘Chronic exposure to malaria is associated with inhibitory and activation markers on atypical memory B cells and marginal zone-like B cells’, Frontiers in Immunology, 8(AUG), pp. 1–14. Available at: 10.3389/fimmu.2017.00966.

53. Venkatraman, N. et al. (2025) ‘Evaluation of a novel malaria anti-sporozoite vaccine candidate, R21 in Matrix-M adjuvant, in the UK and Burkina Faso: two phase 1, first-in-human trials’, The Lancet Microbe, 6(3). Available at: 10.1016/S2666-5247(24)00084-3.

54. White, M.T. et al. (2014) ‘A combined analysis of immunogenicity, antibody kinetics and vaccine efficacy from phase 2 trials of the RTS,S malaria vaccine’, BMC Medicine, 12(1). Available at: 10.1186/s12916-014-0117-2.

55. WHO (2021) WHO recommends groundbreaking malaria vaccine for children at risk, World Health Organization. Available at: https://www.who.int/news/item/06-10-2021-who-recommends-groundbreaking-malaria-vaccine-for-children-at-risk#:~:text=The

55a. World Health Organization (WHO,falciparum malaria transmission.

56. WHO (2023a) WHO recommends R21/Matrix-M vaccine for malaria prevention in updated advice on immunization, World Health Organization. Available at: https://www.who.int/news/item/02-10-2023-who-recommends-r21-matrix-m-vaccine-for-malaria-prevention-in-updated-advice-on-immunization (Accessed: 6 October 2023).

57. WHO (2023b) World Malaria Report 2023. World Health Organization.

58. Williams, B.G. et al. (2024) ‘Development of an improved blood-stage malaria vaccine targeting the essential RH5-CyRPA-RIPR invasion complex’, Nature Communications, 15(1). Available at: 10.1038/s41467-024-48721-3.

59. Yu, D. et al. (2022) ‘Targeting TFH cells in human diseases and vaccination: rationale and practice’, Nature Immunology, 23(8), pp. 1157–1168. Available at: 10.1038/s41590-022-01253-8.

60. Zimmermann, P. and Curtis, N. (2019) ‘Factors that influence the immune response to vaccination’, Clinical Microbiology Reviews. Available at: 10.1128/CMR.00084-18.

