## Supplementary for "B and T cell responses to pre-erythrocytic R21/Matrix-M and blood-stage RH5.1/Matrix-M malaria vaccines in endemic settings"

Supplementary Materials.

**Supplementary Table 1: Sample size per assay per timepoint (as determined by sample availability).**

| **R21/Matrix-M**^®^ **(NCT03580824)** | | | | |
| --- | --- | --- | --- | --- |
| Assay | Group 1A/B | Group 3A/C | Group 3B/D | Group 3E |
| NANP-specific B cells | D0: n=4  V3+28: n= 18 | D0: n=5  V3+28: n= 18 | D0: n=4  V3+28: n=18 | D0: n=5  V3+28: n= 15 |
| *Ex vivo* total CD4+ and cTfh | D0: n=12  V3+7: n=18 | D0: n=7  V3+7: n=18 | D0: n=4  V3+28: n=18 | D0: n=2  V3+7: n=18 |
| **RH5.1/Matrix-M**^®^ **(NCT04318002)** | | | | |
| Assay | Group 2A | Group 2B | Group 2C | Group 2D |
| RH5-specific B cells | D0: n=11  V3+28: n=12 | D0: n=10  V3+28: n= 11 | D0: n=8  V3+28: n= 12 | D0: n=5  V3+28: n=10 |
| AIM (RH5-specific CD4+ and cTfh cells) | D0: n=9  V3+7: n=12 | D0: n=7  V3+7: n=11 | D0: n=9  V3+7: n=12 | D0: n=9  V3+28: n=11 |
| **RH5.1/AS01_B_ (NCT02927145)** | | | | |
| Assay | Group 2 | Group 3 |  |  |
| RH5-specific B cells | V3+28: n=3 | V3+28: n=6 |  |  |

Day 0 (D0): baseline, V3: Vaccination 3, V3+7: 7 days post-third vaccination (V3), V3+28: 28 days post-third vaccination (V3). AIM = Activation Induced Marker Assay.

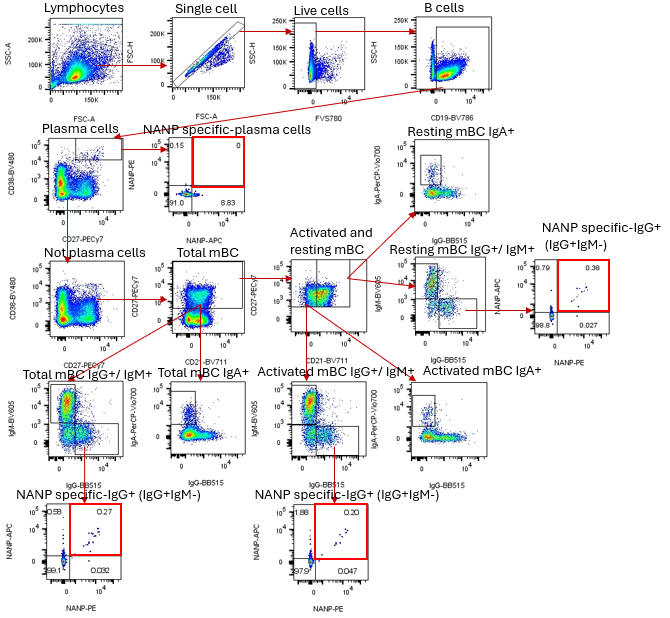

**Supplementary Figure 1. Representative gating strategy to identify antigen-specific plasma and memory B cells.**

Cryopreserved PBMCs from Day 0 (D0) and V3+28 (28 days post-third vaccination (V3)) were stained *ex vivo* and analysed by flow cytometry. Antigen-specific cells within plasma and memory B cell populations were defined as follows. Lymphocytes were defined by forward scatter area (FSC-A) and side scatter area (SSC-A), and then single cells were selected based on FSC height (FSC-H) versus FSC-A, which excludes aggregated cells. Next, live cells were defined as negative for viability stain FVS780. Total B cells were identified as CD19+. Within live B cells, plasma cells were identified as CD38++CD27+. Antigen-specific plasma cells were defined based on co-staining with both PE and APC vaccine-specific probes (example of NANP-specific plasma cells shown with red outline as NANP-PE+/NANP-APC+). Using a “NOT” gate, non-plasma cells were further analysed based on exclusion of the plasma cell population from total live B cells. Within the non-plasma cells, total memory B cells were identified as CD27+, while resting and activated memory B cells were defined identified as CD27+CD21+ and CD27-CD21+, respectively. Memory subsets were further classified as IgG+ (IgG+IgM-), IgM+ (IgG-IgM+), or IgA+ (IgA+IgG-). Antigen-specific memory B cells were identified by probe staining as per plasma cells analyses (examples of NANP-specific cells shown with red outline as NANP-PE+/NANP-APC+ within total, resting, and activated memory IgG+ cells).

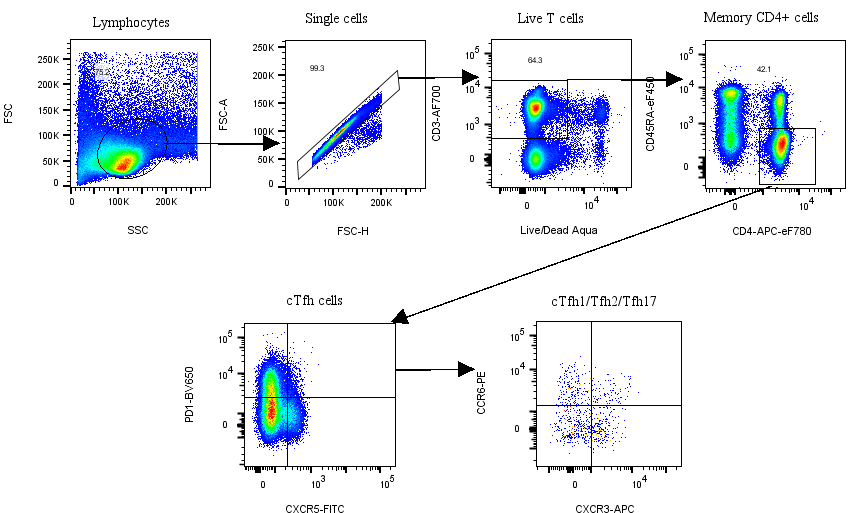

**Supplementary Figure 2. Representative gating strategy to define total memory CD4+ T cells and circulating Tfh (cTfh) cells for R21/Matrix-M**^®^ **analyses.**

Cryopreserved PBMCs from Day 0 ((D0); baseline) and V3+7 (7 days post-third vaccination (V3)) were stained *ex vivo* and analysed by flow cytometry. Memory CD4+ and circulating Tfh (cTfh) cells were defined as follows. Lymphocytes were defined by forward scatter area (FSC-A) and side scatter area (SSC-A), and then single cells were selected based on FSC height (FSC-H) versus FSC-A, which excludes aggregated cells. Next, live T cells were defined as CD3+ and negative for viability stain live/dead Aqua. Memory CD4+ cells were identified as CD4+CD45RA-, and memory cTfh cells were defined as CXCR5+PD1+ within these memory CD4+ T cells.

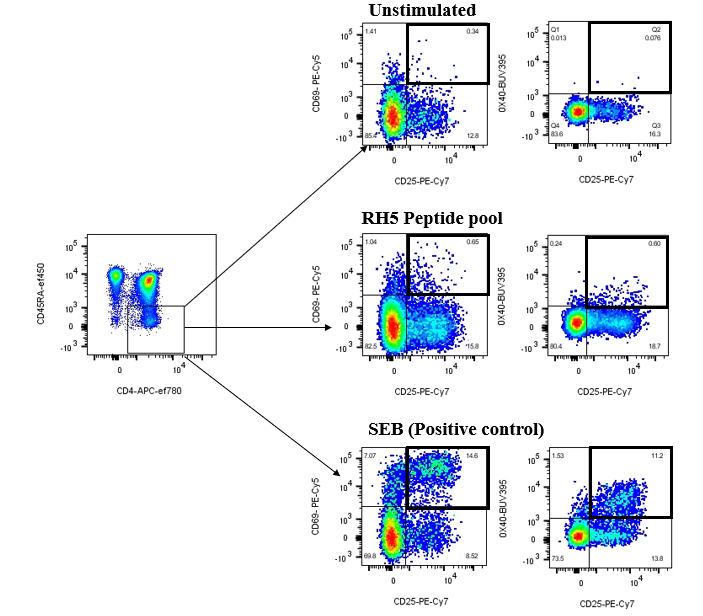

**Supplementary Figure 3. Representative gating strategy to identify RH5-specific memory CD4+ T cells.**

Cryopreserved PBMCs from Day 0 ((D0); baseline) and V3+7 (7 days post-third vaccination (V3)) were stimulated with medium alone, an RH5 peptide pool, or an SEB positive control for 24h, then stained and analysed. The total memory CD4+ T population was defined as CD4+CD45RA- shown in Supplementary Figure 2. Within these, RH5-specific memory CD4+ T cells were identified using an “OR” gate based on co-expression of CD25 with CD69 and/or CD25 with OX40 (shown in thick black boxes) in response to stimulation with the RH5 peptide pool. Frequencies of RH5-specific cells were reported following background subtraction of frequencies of CD25+CD69+ and/or CD25+OX40+ cells in the unstimulated well from matched samples.

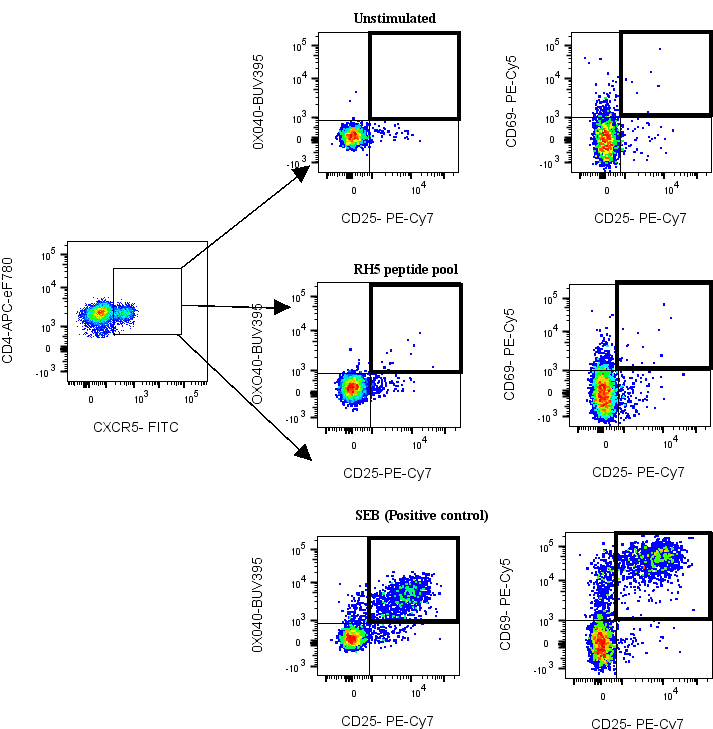

**Supplementary Figure 4. Representative gating strategy to identify RH5-specific circulating memory Tfh (cTfh) cells.**

Cryopreserved PBMCs from Day 0 ((D0); baseline) and V3+7 (7 days post-third vaccination (V3)) were stimulated with medium alone, an RH5 peptide pool, or an SEB positive control for 24h, then stained and analysed. The total memory CXCR5+ cTfh population and cTfh1/2/17 subsets were identified as shown in Supplementary Figure 2. Within these, RH5-specific memory cTfh cells were identified using an “OR” gate based on co-expression of CD25 with CD69 and/or CD25 with OX40 (shown in thick black boxes) in response to stimulation with the RH5 peptide pool. Frequencies of RH5-specific cells were reported following background subtraction of frequencies of CD25+CD69+ and/or CD25+OX40+ cells in the unstimulated well from matched samples.

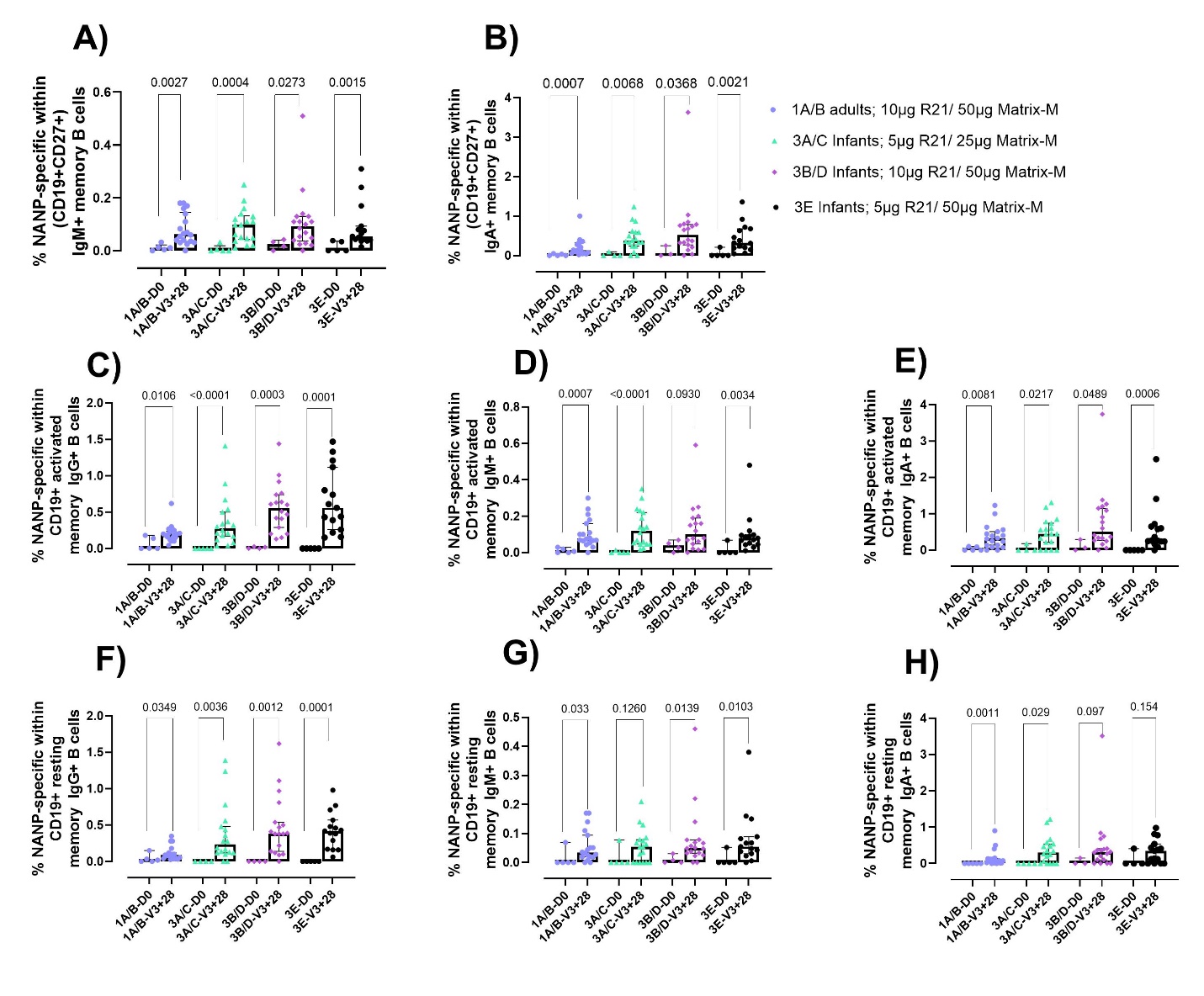

**Supplementary Figure 5. Total/ activated/ resting NANP-specific memory B cells by isotype.**

Cryopreserved PBMCs from Day 0 (D0) and V3+28 (28 days post-third vaccination (V3)) were stained *ex vivo* and analysed by flow cytometry. NANP-specific cells within total/ activated/ resting memory B cell populations were defined as shown in Figure 1 and Supplementary Figure 1. Frequencies of NANP-specific were compared by Mann Whitney tests between (**A**) total IgM+ mBC cells at D0 and V3+28 within groups, (**B**) total IgA+ mBC cells at D0 and V3+28 within groups, (**C**) activated IgG+ mBC cells at D0 and V3+28 within groups, (**D**) activated IgM+ mBC cells at D0 and V3+28 within groups, (**E**) activated IgA+ mBC cells at D0 and V3+28 within groups, (**F**) resting IgG+ mBC cells at D0 and V3+28 within groups, (**G**) resting IgM+ mBC cells at D0 and V3+28 within groups, and (**H**) resting IgA+ mBC cells at D0 and V3+28 within groups. Group 1A/B: 10μg R21/50μg Matrix-M^®^ (adults); Group 3A/C: 5μg R21/25μg Matrix-M^®^ (infants), Group 3B/D: 10μg R21/50μg Matrix-M^®^ (infants); and, Group 3E: 5μg R21/50μg Matrix-M^®^ (infants). Sample sizes for all assays were based on sample availability; each point represents a single sample. Bars and error bars represent medians and interquartile ranges, respectively. Significant p values are annotated on graphs; p < 0.05 was considered significant. mBC = memory B cells.

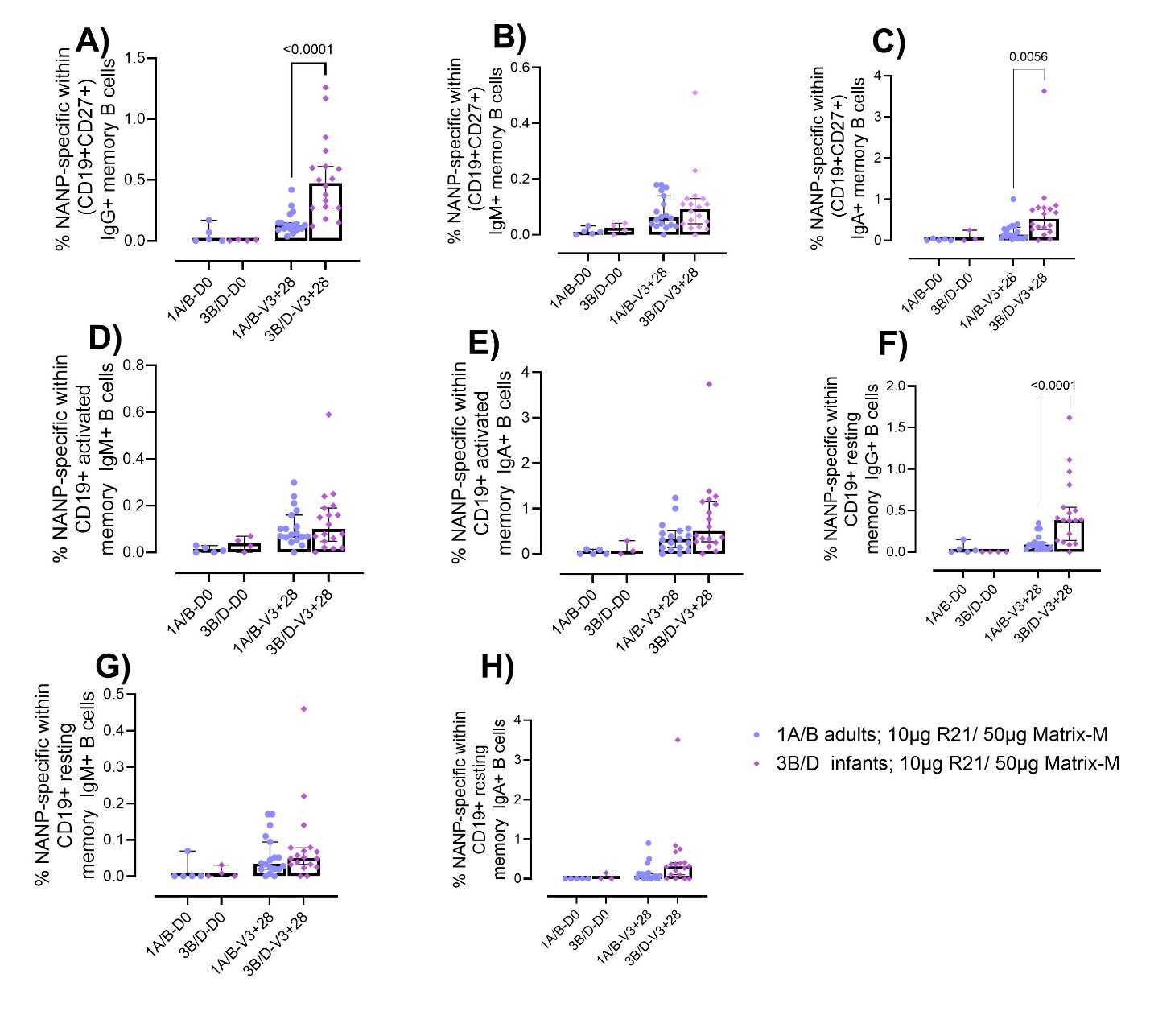

**Supplementary Figure 6. Comparison of NANP-specific memory B cell responses by age.**

Cryopreserved PBMCs from Day 0 (D0) and V3+28 (28 days post-third vaccination (V3)) were stained *ex vivo* and analysed by flow cytometry. NANP-specific cells within total/ activated/ resting memory B cell populations were defined as shown in Figure 1 and Supplementary Figure 1. Frequencies of NANP-specific cells at V3+28 were compared between adult (1A/B) and infant (3B/D) groups receiving the same 10μg R21/ 50μg Matrix-M^®^ dose by Mann Whitney test within (**A**) total IgG+ memory B cells, (**B**) total IgA+ memory B cells, and (**C**) total IgM+ memory B cells, (**D**) activated IgM+ memory B cells, (**E**) activated IgA+ memory B cells, (**F**) resting IgG+ memory B cells, (**G**) resting IgA+ memory B cells, and (**H**) resting IgM+ memory B cells. Sample sizes for all assays were based on sample availability; each point represents a single sample. Bars and error bars represent medians and interquartile ranges, respectively. Significant p values are annotated on graphs; p < 0.05 was considered significant.

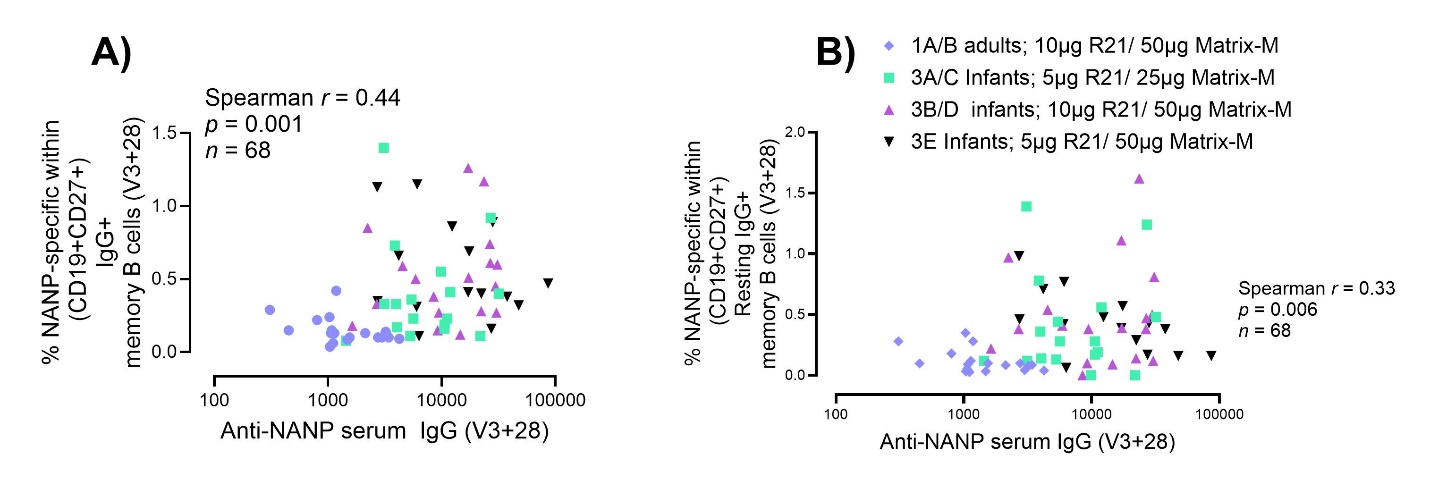

**Supplementary Figure 7. Correlations of NANP-specific IgG+ total and resting memory B cells against anti-NANP serum IgG.**

Spearman correlations were performed between anti-NANP serum IgG V3+28 (28 days post-third vaccination (V3)) and (**A**) NANP-specific total IgG+ memory B cells, and (**B**) NANP-specific resting IgG+ memory B cells frequencies, both also measured at V3+28.

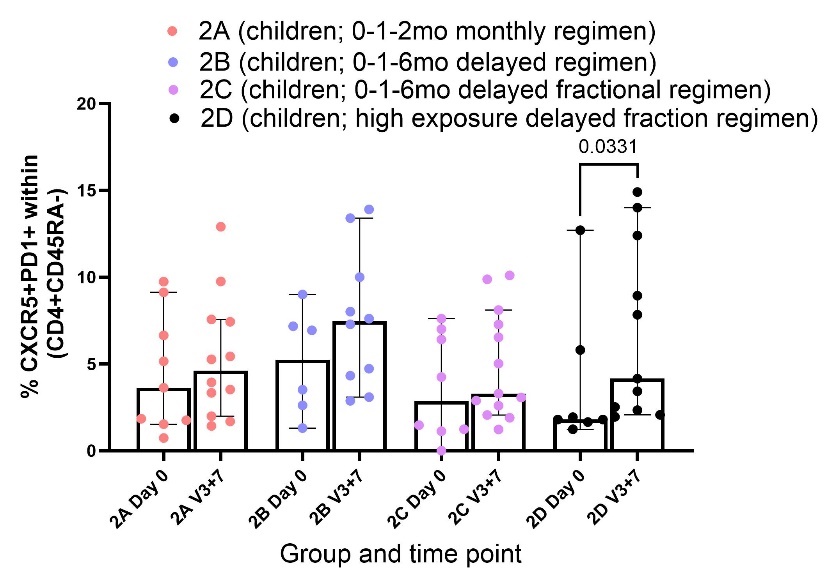

**Supplementary Figure 8. Frequencies of CXCR5+PD1+ circulating Tfh (cTfh) cells in RH5.1/Matrix-M**^®^ **vaccinees.**

Cryopreserved PBMCs from Day 0 ((D0) (baseline) and V3+7 (7 days post-third vaccination (V3)) were incubated with medium alone for 24h, then stained and analysed. The total memory CD4+ T population was identified as shown in Supplementary Figure 2, then CXCR5+PD1+ cTfh cells were identified within the total memory CD4+ T cell population (gating shown in Supplementary Figure 2) and compared between D0 and V3+7 within each group by Mann-Whitney tests. Sample sizes for all assays were based on sample availability; each point represents a single sample. Bars and error bars represent medians and interquartile ranges, respectively. Significant p values are annotated on graphs (none); p < 0.05 was considered significant.

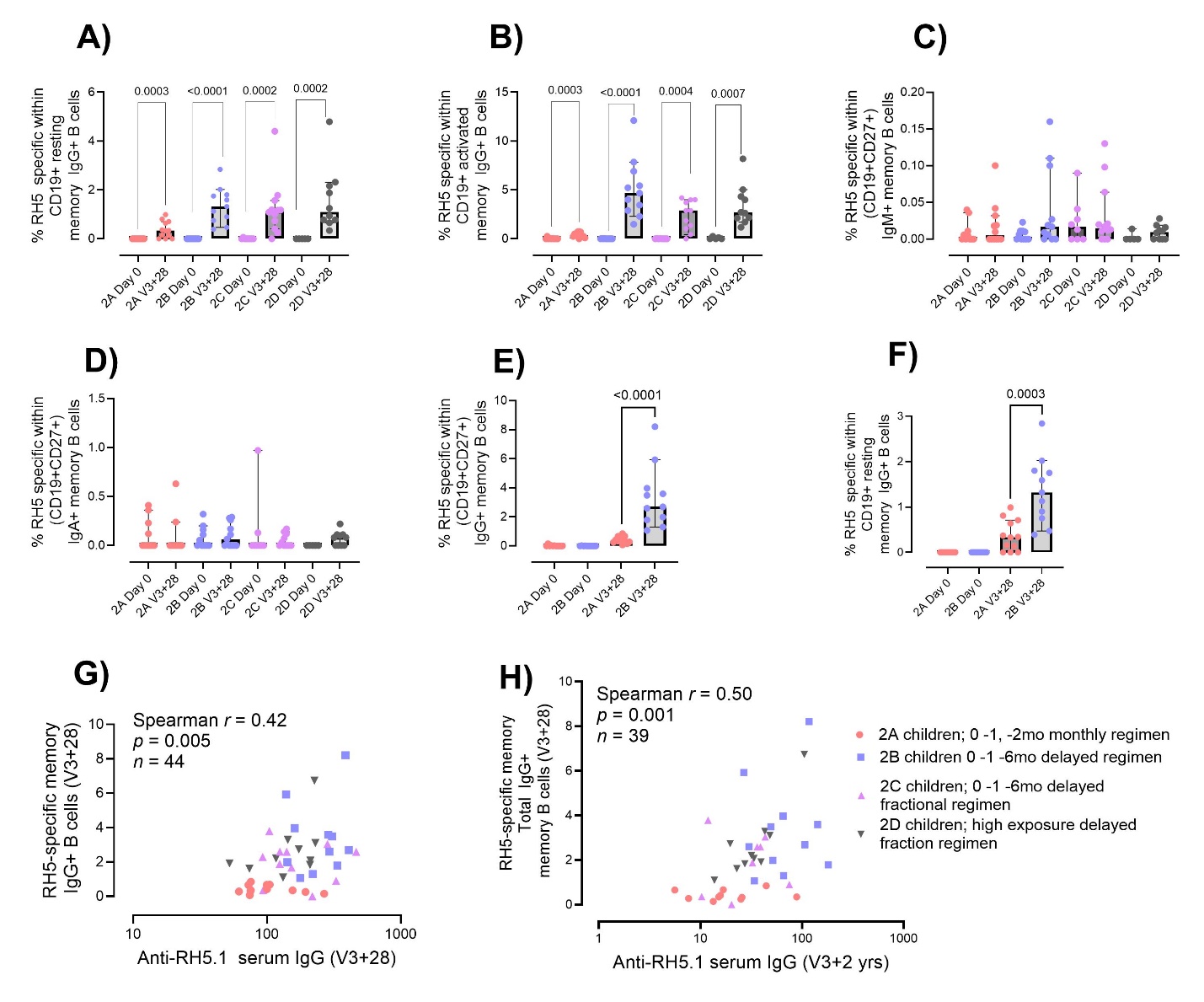

**Supplementary Figure 9. RH5-specific memory B cells and correlations with anti-RH5.1 serum IgG.**

Cryopreserved PBMCs from Day 0 (D0) and V3+28 (28 days post-third vaccination (V3)) were stained *ex vivo* and analysed by flow cytometry. RH5-specific memory B cell subsets were defined as shown in Figure 3 and Supplementary Figure 1. Frequencies were compared between D0 and V3+28 within groups by Mann Whitney test within (**A**) resting IgG+ memory B cells, (**B**) activated IgG+ memory B cells, (**C**) total IgM+ memory B cells, and (**D**) total IgA+ memory B cells. Frequencies of RH5-specific cells were also compared between “monthly” (Group 2A) vs “delayed” (Group 2B) groups within (**E**) total IgG+ memory B cells, and (**F**) resting IgG+ memory B cells. Spearman correlation analyses was performed between RH5-specific total IgG+ memory B cells and anti-RH5.1 serum IgG at (**G**) V3+28 and (**H**) V3+2yr. Sample sizes for all assays were based on sample availability; each point represents a single sample. Bars and error bars represent medians and interquartile ranges, respectively. Significant p values are annotated on graphs; p < 0.05 was considered significant.
